# Loss-of-function of *MFGE8* and protection against coronary atherosclerosis

**DOI:** 10.1101/2021.06.23.21259381

**Authors:** Sanni E. Ruotsalainen, Ida Surakka, Nina Mars, Juha Karjalainen, Mitja Kurki, Masahiro Kanai, Kristi Krebs, Pashupati P. Mishra, Binisha H. Mishra, Juha Sinisalo, Priit Palta, Terho Lehtimäki, Olli Raitakari, Estonian Biobank research team, Lili Milani, The Biobank Japan Project, Yukinori Okada, FinnGen, Aarno Palotie, Elisabeth Widen, Mark J. Daly, Samuli Ripatti

## Abstract

Cardiovascular diseases are the leading cause of premature death and disability worldwide, with both genetic and environmental determinants. While genome-wide association studies have identified multiple genetic loci associated with cardiovascular diseases, exact genes driving these associations remain mostly uncovered. Due to Finland’s population history, many deleterious and high-impact variants are enriched in the Finnish population giving a possibility to find genetic associations for protein-truncating variants that likely tie the association to a gene and that would not be detected elsewhere.

In FinnGen, a large Finnish biobank study, we identified an inframe insertion rs534125149 in *MFGE8* to have protective effect against coronary atherosclerosis (OR = 0.75, p = 2.63×10^-16^) and related endpoints. This variant is highly enriched in Finland (70-fold compared to Non-Finnish Europeans) with allele frequency of 3% in Finland. The protective association was replicated in meta-analysis of biobanks of Japan and Estonian (OR = 0.75, p = 5.41×10^-7^).

Additionally, we identified a splice acceptor variant rs201988637 in *MFGE8*, independent of the rs534125149 and similarly protective in relation to coronary atherosclerosis (OR = 0.72, p = 7.94×10^-06^) and related endpoints, with no significant risk-increasing associations. The protein-truncating variant was also associated with lower pulse pressure, pointing towards a function of *MFGE8* in arterial stiffness and aging also in humans in addition to previous evidence in mice. In conclusion, our results show that inhibiting the production of lactadherin could lower the risk for coronary heart disease substantially.

## Introduction

Cardiovascular disease (CVD) is the leading cause of premature death and disability worldwide, with both genetic and environmental determinants^1, 2^. The most common cardiovascular disease is coronary heart disease (CHD), including coronary atherosclerosis and myocardial infarction, among others. While genome-wide association studies (GWAS) have identified multiple genetic loci associated with cardiovascular diseases, exact genes driving these associations remain mostly uncovered^3^.

Due to Finland’s population history, many deleterious and high-impact variants are enriched in the Finnish population giving a possibility to find genetic associations that would not be detected elsewhere^4^. Many studies have reported high-impact loss-of-function (LoF) variants associated with risk factors for CVD, such as blood lipid levels, thus impacting on the CVD risk remarkably. For example, high-impact LoF variants in genes *LPA*^4^*, PCSK9*^5^*, APOC3*^6^ and *ANGPTL4* ^7^ have been shown to be associated with Lipoprotein(a), LDL-cholesterol (LDL-C) or triglyceride levels and lowering the CVD risk.

Besides blood lipids, other risk factors for CVD include hypertension, smoking and the metabolic syndrome cluster components. The mechanism that links these risk factors to atherogenesis, however, remains incompletely elucidated. Many, if not all, of these risk factors, however, also participate in the activation of inflammatory pathways, and inflammation in turn can alter the function of artery wall cells in a manner that drives atherosclerosis^8^.

Using data from a sizeable Finnish biobank study FinnGen (n = 260 405), we identified an inframe insertion rs534125149 in *MFGE8* to protect against coronary atherosclerosis and related endpoints, such as myocardial infarction (MI). This variant is highly enriched in Finland, 70- fold compared to Non-Finnish Europeans (NFE) in the gnomAD genome reference database^9^ with AF of 3% in Finland. This association was also replicated in BioBank Japan (BBJ) and Estonian Biobank (EstBB). We also identified a splice acceptor variant rs201988637 in the same gene, which is also protective against coronary atherosclerosis-related endpoints, indicating that rs534125149 act as a loss-of-function variant in *MFGE8*. Associations of functional variants in *MFGE8* were specific to coronary atherosclerosis-related endpoints, and they did not significantly (p < 1.75×10^-5^) increase risk for any other disease, highlighting *MFGE8* as a potential drug target candidate.

## Material and methods

### Study cohort and data

We studied total of 2 861 disease endpoints in Finnish biobank study FinnGen (n = 260 405) (**Table 1**). FinnGen (https://www.finngen.fi/en) is a large biobank study that aims to genotype 500 000 Finns and combine this data with longitudinal registry data, including national hospital discharge, death, and medication reimbursement registries, using unique national personal identification numbers. FinnGen includes prospective epidemiological and disease-based cohorts as well as hospital biobank samples.

**Table 1:** Basic characteristics of the study cohort.

|  | All | Females | Males |
| --- | --- | --- | --- |
| N (%) | 260 405 | 147 061 (56.47 %) | 113 344 (43.53 %) |
| Age (mean (sd)) | 53.15 (17.55) | 51.84 (17.71) | 54.85 (17.19) |
| BMI (mean (sd)) <sup>a</sup> | 27.29 (5.36) | 27.21 (5.83) | 27.38 (4.76) |
| Statin use (N (%)) | 86 466 (33.2 %) | 40 422 (27.48 %) | 46 044 (40.62 %) |
| Hypertension (N (%)) | 68 005 (26.11 %) | 33 420 (22.72 %) | 34 585 (30.51 %) |
| Smoking (N (%)) <sup>b</sup> | 1 733 (1.07 %) | 901 (0.96 %) | 832 (1.22 %) |
| Coronary atherosclerosis | 28 598 (11.38 %) | 9 252 (6.87 %) | 19 346 (17.86 %) |
| Myocardial infarction | 14 305 (6.04 %) | 3 958 (2.87 %) | 10 347 (10.42 %) |

<sup>a</sup> BMI is available only from 178 966 individuals
<sup>b</sup> Smoking information is available only from 98 654 individuals

### Definition of disease endpoints

All the 2 861 disease-endpoint analysed in FinnGen have been defined based on registry linkage to national hospital discharge, death, and medication reimbursement registries. Diagnoses are based on International Classification of Diseases (ICD) codes and have been harmonized over ICD codes 8, 9 and 10. More detailed lists of the ICD codes used for the disease-endpoints myocardial infarction and coronary atherosclerosis, which are discussed more in this study, are in Supplementary Material. A complete list of endpoints analysed, and their definitions is available at https://www.finngen.fi/en/researchers/clinical-endpoints.

### Genotyping and imputation

FinnGen samples were genotyped with multiple Illumina and Affymetrix arrays (Thermo Fisher Scientific, Santa Clara, CA, USA). Genotype calls were made with GenCall and zCall algorithms for Illumina and AxiomGT1 algorithm for Affymetrix chip genotyping data batchwise. Genotyping data produced with previous chip platforms were lifted over to build version 38 (GRCh38/hg38) following the protocol described here: dx.doi.org/10.17504/protocols.io.nqtddwn. Samples with sex discrepancies, high genotype missingness (> 5%), excess heterozygosity (±4SD) and non-Finnish ancestry were removed. Variants with high missingness (> 2%), deviation from Hardy–Weinberg equilibrium (P < 1×10^-6^) and low minor allele count (MAC < 3) were removed.

Pre-phasing of genotyped data was performed with Eagle 2.3.5 (https://data.broadinstitute.org/alkesgroup/Eagle/) with the default parameters, except the number of conditioning haplotypes was set to 20,000. Imputation of the genotypes was carried out by using the population-specific Sequencing Initiative Suomi (SISu) v3 imputation reference panel with Beagle 4.1 (version 08Jun17.d8b, https://faculty.washington.edu/browning/beagle/b4_1.html) as described in the following protocol: dx.doi.org/10.17504/protocols.io.nmndc5e. SISu v3 imputation reference panel was developed using the high-coverage (25–30x) whole-genome sequencing data generated at the Broad Institute of MIT and Harvard and at the McDonnell Genome Institute at Washington University, USA; and jointly processed at the Broad Institute. Variant callset was produced with Genomic Analysis Toolkit (GATK) HaplotypeCaller algorithm by following GATK best practices for variant calling. Genotype-, sample- and variant-wise quality control was applied in an iterative manner by using the Hail framework v0.2. The resulting high-quality WGS data for 3 775 individuals were phased with Eagle 2.3.5 as described above. As a post-imputation quality control, variants with INFO score < 0.7 were excluded.

### Association testing

A total of 260 405 samples from FinnGen Data Freeze 6 with 2 861 disease endpoints were analyzed using Scalable and Accurate Implementation of Generalized mixed model (SAIGE), which uses saddlepoint approximation (SPA) to calibrate unbalanced case-control ratios^10^. Models were adjusted for age, sex, genotyping batch and first ten principal components. All variants reaching genome-wide significance p-value threshold of 5×10^-8^ are considered as genome-wide significant (GWS), and all disease- endpoints reaching multiple testing corrected (for the number of endpoints tested = 2 861) p-value threshold of 0.05/2 861 = 1.75×10^-5^ were considered as phenome-wide significant (PWS).

Independent GWS loci for atherosclerosis were determined as adding ±0.5Mb around each variant that reach the genome-wide significance threshold, overlapping regions were merged. The NHGRI- EBI GWAS Catalog^11^ was used for assessing the novelty of the independent loci with any CVD- related endpoint or traditional risk factor for CVD, such as blood lipids, BMI and blood pressure. All novel loci for CVD were fine-mapped using FINEMAP^12^ to determine the credible sets in each signal.

In Corogene^13^ (n = 5 300), a sub-cohort of FinnGen where participants have been collected as patients with coronary artery disease CAD) and other related heart diseases, we tested the association of rs534125149 with sub-types of coronary heart disease (acute coronary syndrome, stable coronary heart disease (CHD) and other heart attacks). The acute coronary syndrome was further divided into unstable Angina pectoris, non-ST segment elevation myocardial infarction (NSTEMI) and ST- segment elevation myocardial infarction (STEMI). Associations were tested by calculating risk ratios (RR) for carriers vs. non-carriers of rs534125149 using non-CHD group always as controls and excluding the other tested groups from the analysis. P-values were calculated using χ^2^- test, and p- values < 0.05 were considered significant.

### Survival analysis

Survival analysis for coronary atherosclerosis and myocardial was performed using GATE^14^, which accounts for both population structure and sample relatedness and controls type I error rates even for phenotypes with extremely heavy censoring. GATE transforms the likelihood of a multivariate Gaussian frailty model to a modified Poisson generalized linear mixed model (GLMM^15, 16^), and to obtain well-calibrated p-values for heavily censored phenotypes, GATE uses the SPA to estimate the null distribution of the score statistic. For coronary atherosclerosis and myocardial infarction, survival time from birth to first diagnose was analyzed for both rs534125149 and rs201988637. Models were adjusted for age, sex, genotyping batch and first ten principal components, similarly to original GWAS analyses.

### Replication and biomarker analyses

We tested the association of the two *MFGE8* variants (rs534125149 and rs201988637) with quantitative measurements of cardiometabolic relevance or known risk factors for CVD in two sub-cohorts of FinnGen, the population-based national FINRISK study^17^ (n = 26 717) and GeneRISK^18^ (n = 7 239). The associations were tested across 66 quantitative measurements of cardiometabolic relevance in FINRISK, and for 158 sub-lipid species in GeneRISK. In Young Finns Study (YFS)^19^ cohort (n = 1 934), we tested the association of the two variants with three measurements of arterial relevance (carotid artery distensibility, pulse wave velocity and pulse pressure).

In addition to Finnish cohorts described above, we tested the association of the two variants in Estonian Biobank data (EstBB)^20, 21^, BioBank Japan (BBJ) ^22, 23^ and UK Biobank (UKBB)^24^. In EstBB (n = 51 388-137 722) we tested the associations of both variants with body mass index (BMI), systolic and diastolic blood pressure (SBP and DBP) and pulse pressure (PP), in BBJ in we tested the association of rs534125149 with 17 known quantitative risk factors for CVD and lastly, in the UKBB we tested the association of rs201988637 with 79 measurements of cardiometabolic relevance. In all of these biomarker analyses, a linear regression model adjusted for age and sex was used and for all quantitative risk factors rank-based inverse normal transformation was applied prior to analysis. Bonferroni corrected p-value threshold for the number of phenotypes tested was used to assess the significance of resulting associations in each cohort.

For biomarkers that showed significant association in any of the cohorts, we performed a meta-analysis across all cohorts the measurement was available. Meta-analysis was performed using inverse-variance weighted fixed-effects meta-analysis method^25, 26^. Bonferroni corrected p-value for number of traits tested (n = 2) was used to assess the significance of resulting associations in meta-analysis.

### Identifying causal variants

We used FINEMAP^12^ on the GWAS summary statistics to identify causal variants underlying the associations for MI (strict definition, i.e., only primary diagnoses accepted) and coronary atherosclerosis. FINEMAP analyses were restricted to a ±1.5Mb region around the rs534125149. We assessed variants in the top 95% credible sets, i.e., the sets of variants encompassing at least 95% of the probability of being causal (causal probability) within each causal signal in the genomic region. Credible sets were filtered if minimum linkage disequilibrium (LD, r^2^) between the variants in the credible set was < 0.1, i.e., not clearly representing one signal.

## Results

### GWAS results for Coronary Atherosclerosis

We identified a total of 2 302 variants associated (GWS, p < 5×10^-8^) with coronary atherosclerosis (detailed description of the definition of the endpoint is in Supplementary Material). These variants were located in 38 distinct genetic loci (a minimum of 0.5 Mb distance from each other; **Figure 1**). Out of the 38 GWS loci, four (within or near genes *MFGE8, TMEM200A, PRG3* and *FHL1*) were novel for any CVD-related endpoints or risk factor for CVD compared to the GWAS Catalog^11^ [https://www.ebi.ac.uk/gwas/] (**Supplementary Table 1**). Lead variants in these novel loci and their characteristics are listed in **Table 2** and locus zoom plots for each of the loci are in **Supplementary Figure 1**.

**Figure 1:**
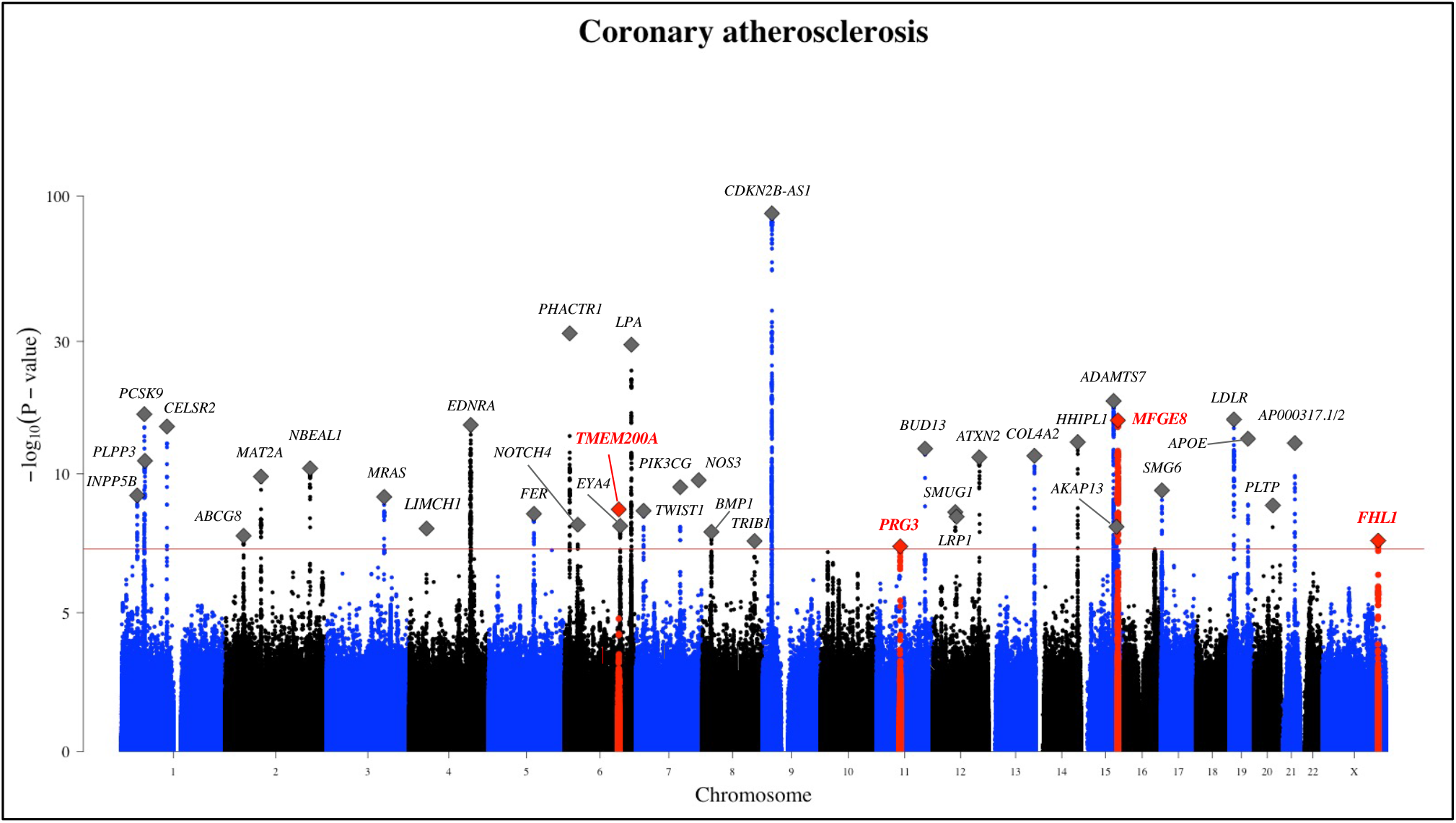
GWAS results for coronary atherosclerosis in FinnGen. Total number of independent genome-wide significant associations (GWS; p < 5×10^-8^) is 38, the lead variant in each marked with diamonds. Four novel associations for CVD- related phenotypes are highlighted with ±750 Mb around the lead variant in the region as red and the lead variant marked with red diamond.

**Table 2:**
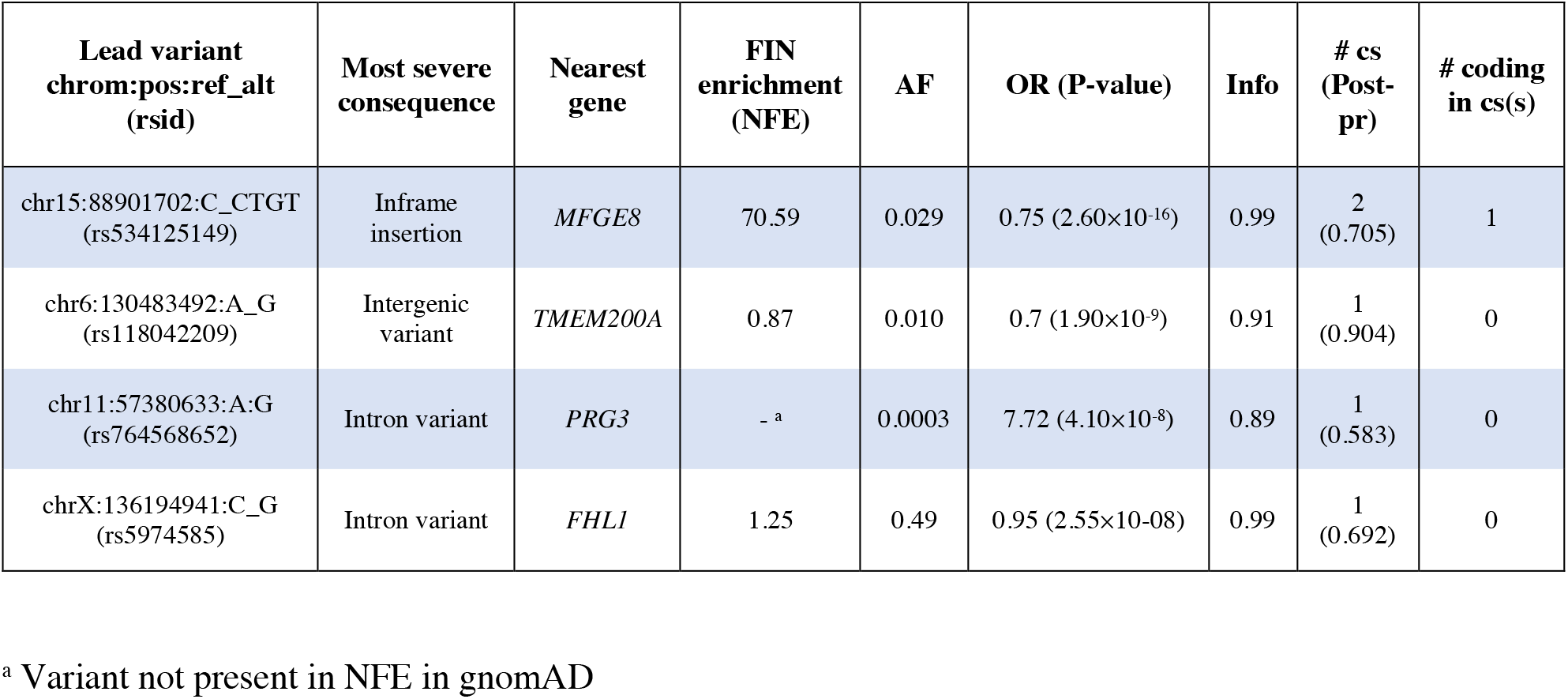
Lead variants in novel loci associated with coronary atherosclerosis.

| Lead variant<br>chrom:pos:ref_alt<br>(rsid) | Most severe<br>consequence | Nearest<br>gene | FIN<br>enrichment<br>(NFE) | AF | OR (P-value) | Info | # cs<br>(Post-<br>pr) | # coding<br>in cs(s) |
| --- | --- | --- | --- | --- | --- | --- | --- | --- |
| chr15:88901702:C_CTGT<br>(rs534125149) | Inframe<br>insertion | *MFGE8* | 70.59 | 0.029 | 0.75 (2.60×10<sup>-16</sup>) | 0.99 | 2<br>(0.705) | 1 |
| chr6:130483492:A_G<br>(rs118042209) | Intergenic<br>variant | *TMEM200A* | 0.87 | 0.010 | 0.7 (1.90×10<sup>-9</sup>) | 0.91 | 1<br>(0.904) | 0 |
| chr11:57380633:A:G<br>(rs764568652) | Intron variant | *PRG3* | -<sup>a</sup> | 0.0003 | 7.72 (4.10×10<sup>-8</sup>) | 0.89 | 1<br>(0.583) | 0 |
| chrX:136194941:C_G<br>(rs5974585) | Intron variant | *FHL1* | 1.25 | 0.49 | 0.95 (2.55×10<sup>-8</sup>) | 0.99 | 1<br>(0.692) | 0 |

<sup>a</sup> Variant not present in NFE in gnomAD

Among the three novel loci for coronary atherosclerosis, the locus near *MFGE8* had the strongest association (p-value = 2.63×10^-16^ for top variant rs534125149). The lead variant is an inframe insertion located in the sixth exon in the *MFGE8* gene (**Supplementary Figure 2**) and it is highly enriched in the Finnish population compared to NFSEEs (Non-Finnish, Estonian or Swedish Europeans). Interestingly, *MFGE8* is mainly expressed in coronary and tibial arteries according to data from GTEx v8 (**Supplementary Figure 3**).

In addition to *MFGE8*, we identified three additional novel loci to be associated with coronary atherosclerosis, *TMT200A*, *PRG3* and *FHL1* being the nearest genes of the lead variants. *TMT200A* and *PRG3* loci had one non-coding low-frequency variant reaching the genome-wide significance threshold, and *FHL1* had 11. All variants in the credible sets of all these associations were either intergenic or intronic variants and had no reported significant GWAS associations with any trait in the GWAS Catalog or significant eQTL associations in GTEx. The one variant (rs118042209) in the credible set of *TMEM200A* locus was associated with multiple coronary atherosclerosis related endpoints in FinnGen, including coronary atherosclerosis, ischemic heart disease and angina pectoris, whereas the lead variant in the *PRG3* locus was associated with several cardiomyopathy related endpoints. All variants in the credible set of *FHL1* were associated with multiple coronary atherosclerosis and related endpoints in FinnGen, including angina pectoris and ischemic heart disease. *TMEM200A* have been reported to be associated with ten traits (including height and trauma exposure) and *PRG3* with two traits (eosinophil count and eosinophil percentage of white cells) in the GWAS Catalog. *FHL1* gene had no reported associations in GWAS Catalog.

In the *PRG3* locus the lead variant is 40.9 kb away from a variant (chr11:57340143:G:A (rs11603691), the p-value for coronary atherosclerosis risk = 0.0269) which has previously been reported to be associated with HDL- cholesterol (HDL-C). However, our association is independent of the previously reported variant rs11603691 (p-value for rs764568652 in a conditional analysis = 4.36×10^-08^)

### Replication and phenome-wide association results for rs534125149

We observed a highly protective association for the Finnish enriched inframe insertion rs534125149 in the *MFGE8* gene and coronary atherosclerosis related endpoints, including coronary atherosclerosis (OR = 0.72 [0.63-0.83], p = 7.94×10^-06^) and myocardial infarction (MI) (OR = 0.69 [0.58-0.83], p = 9.62×10^-05^) (**Figure 2**). In total, this variant had PWS significant protective effect on 14 disease endpoints, all related to coronary atherosclerosis. The majority of these disease endpoints are highly overlapping and representing major coronary heart disease (CHD) event. For this variant, we did not detect other phenome-wide significant associations among the 2 861 endpoints in our data. Risk-lowering effect of rs534125149 on CHD was replicated in Biobank Japan ^22, 23^ (BBJ) and the Estonian Biobank (EstBB) ^21^ (35 644 cases and 328 461 controls total: OR = 0.754 [0.67-0.84], p = 5.41×10^-7^). Association results for rs534125149 with CHD and MI across different cohorts are shown in **Supplementary Figure 4**.

**Figure 2:**
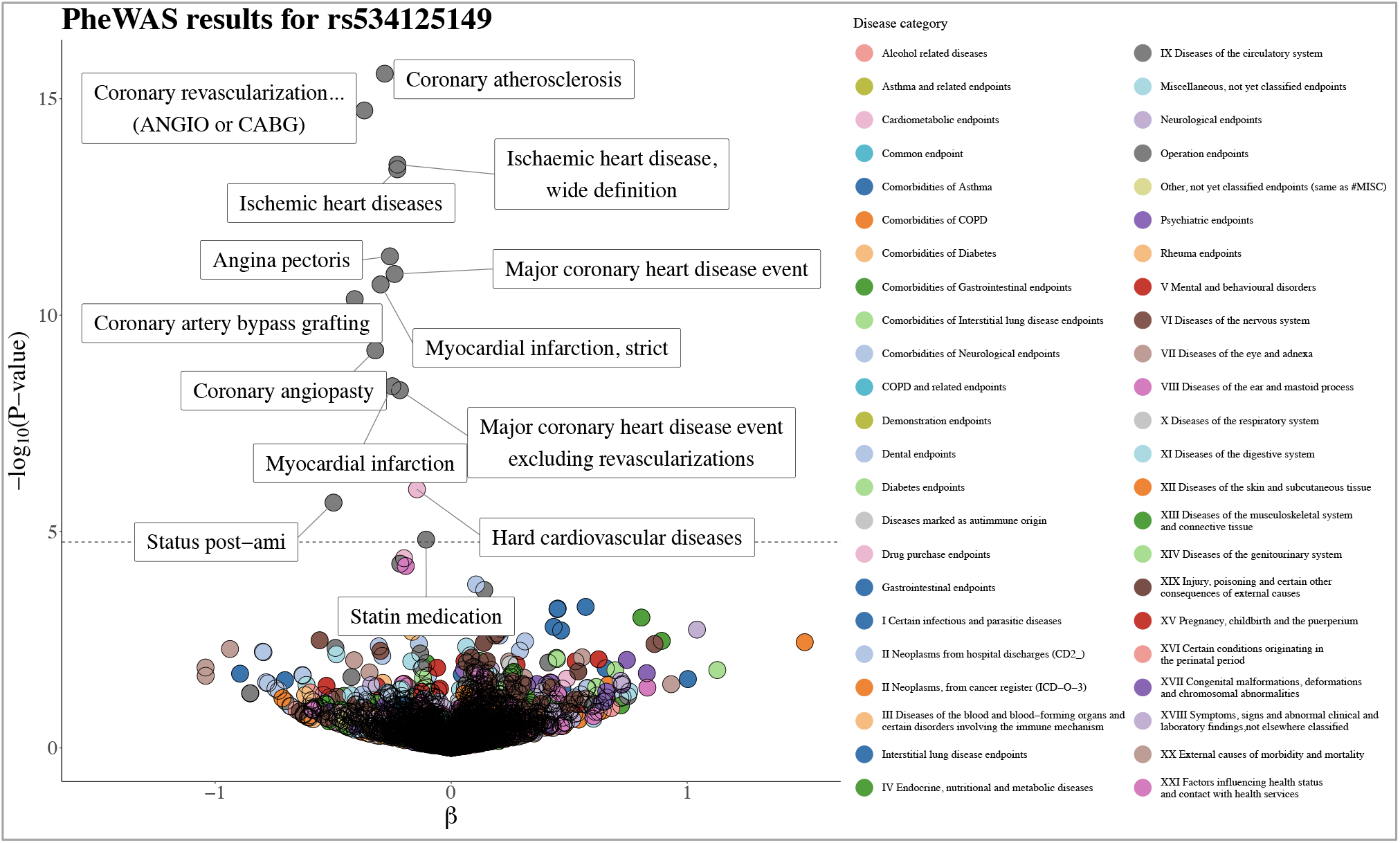
Phenome-wide association study (PheWAS) results for rs534125149. Total number of tested endpoints is 2 861 (A complete list of endpoints analyzed and their definitions is available at https://www.finngen.fi/en/researchers/clinical-endpoints). The dashed line represents the phenome-wide significance threshold, multiple testing corrected by the number of endpoints = 0.05/ 2 861 = 1.75×10^-5^. All endpoints reaching that threshold are labelled in the figure.

### Splice acceptor variant rs201988637 in *MFGE8*

In addition to inframe insertion rs53412514, we also identified a splice acceptor variant (rs201988637) in *MFGE8* to have a protective effect on coronary atherosclerosis (OR = 0.72 [0.63- 0.83], p = 7.94×10^-06^) and related endpoints. The splice acceptor variant had very similar PheWAS profile as the indel (**Supplementary Figure 5**) and furthermore the two variants had very similar protective effect sizes for the endpoints (**Figure 3** and **Supplementary Table 1**). Similar to rs534125149, this variant is also highly enriched in Finland (37- fold compared to NFE), allele frequency in Finland being 0.6 %. Moreover, both the splice acceptor and the inframe insertion variants were enriched to Eastern Finland (**Supplementary Figure 6**).

**Figure 3:**
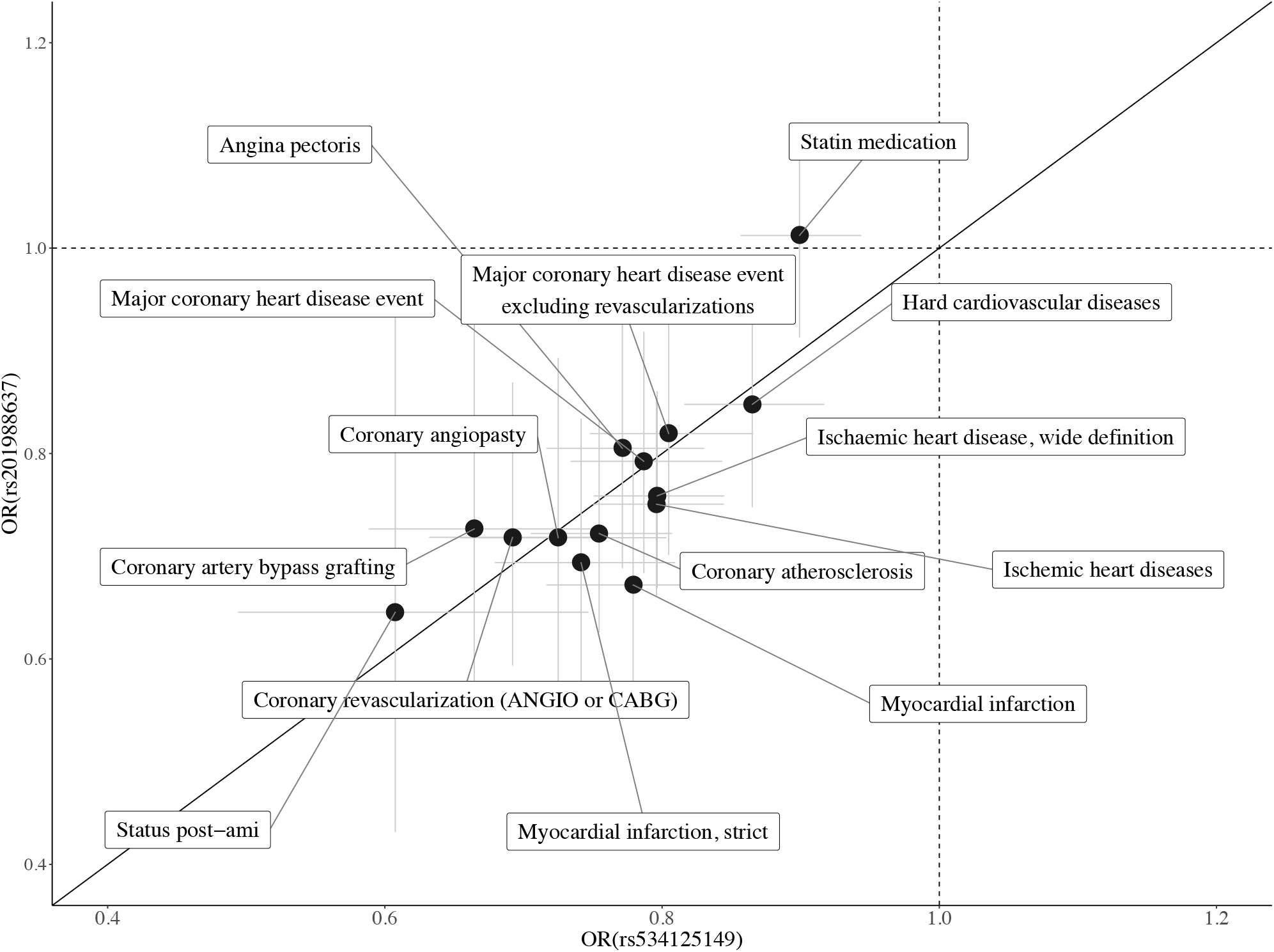
Comparison of the effects (OR) of rs534125149 and rs201988637 for 14 endpoints with p-value < 1.75×10-5 (PWS) for rs534125149. 95% confidence intervals represented as grey lines.

These two variants (rs534125149 and rs201988637) are in low linkage disequilibrium (LD, r^2^ = 0.00015) and did not have any effect on the other variant’s associations with coronary atherosclerosis or MI (**Table 3** and **Supplementary Figure 7**). This indicates that they both have their own, independent protective effect against these endpoints.

**Table 3:** Results of the conditional analysis on MI and coronary atherosclerosis.

| Phenotype | SNPID<br>[chr:position:ref:alt]<br>(rsid) | Most severe<br>consequence | Original GWAS results |  | Conditional results |  |
| --- | --- | --- | --- | --- | --- | --- |
|  |  |  | OR [CI] | P-value | OR [CI] | P-value |
| Coronary<br>atherosclerosis | chr15:88901702:C:CTGT<br>(rs534125149) | Inframe<br>insertion | 0.75 [0.71-0.81] | $2.63 \times 10^{-16}$ | 0.75 [0.70-0.80] <sup>a</sup> | $7.68 \times 10^{-15}$ <sup>a</sup> |
| | chr15:88899813:T:G<br>(rs201988637) | Splice acceptor<br>variant | 0.72 [0.63-0.83] | $7.94 \times 10^{-6}$ | 0.73 [0.64-0.85] <sup>b</sup> | $1.99 \times 10^{-5}$ <sup>b</sup> |
| Myocardial<br>infarction,<br>strict | chr15:88901702:C:CTGT<br>(rs534125149) | Inframe<br>insertion | 0.74 [0.68-0.81] | $1.95 \times 10^{-11}$ | 0.79 [0.73-0.85] <sup>a</sup> | $1.92 \times 10^{-10}$ <sup>a</sup> |
| | chr15:88899813:T:G<br>(rs201988637) | Splice acceptor<br>variant | 0.69 [0.58-0.83] | $9.62 \times 10^{-5}$ | 0.71 [0.59-0.85] <sup>b</sup> | $4.03 \times 10^{-4}$ <sup>b</sup> |

This table present the conditional analysis results for coronary atherosclerosis and MI (strict definition, only primary diagnoses accepted) where the association has been conditioned on rs534125149 and rs201988637 separately
<sup>a</sup> Conditional on rs201988637
<sup>b</sup> Conditional on rs534125149

### Survival analysis

In addition to protection against coronary atherosclerosis and myocardial infarction, both the infame insertion rs534125149 and splice acceptor variant rs201988637 showed also significant association in survival analysis when analyzing survival time from birth to first diagnose of coronary atherosclerosis (HR = 0.78 [0.74-0.93]), p = 1.67×10^-17^ and HR = 0.77 [0.69-0.88], p = 5.08×10^-05^, respectively) and myocardial infarction (HR = 0.86 [0.80-0.93], p = 2.63×10^-10^ and HR = 0.72 [0.61-0.85], p = 8.16×10^-05^). In addition, when combining the heterozygous and homozygous carriers of both rs534125149 and rs201988637 together, carriers get the first diagnose significantly later than non-carriers (HR = 0.81 [0.77-0.85], p = 6.4×10^-16^ for coronary atherosclerosis and HR = 0.78 [0.72-0.85], p = 1.16×10^-11^ for MI) (**Figure 4**).

**Figure 4:**
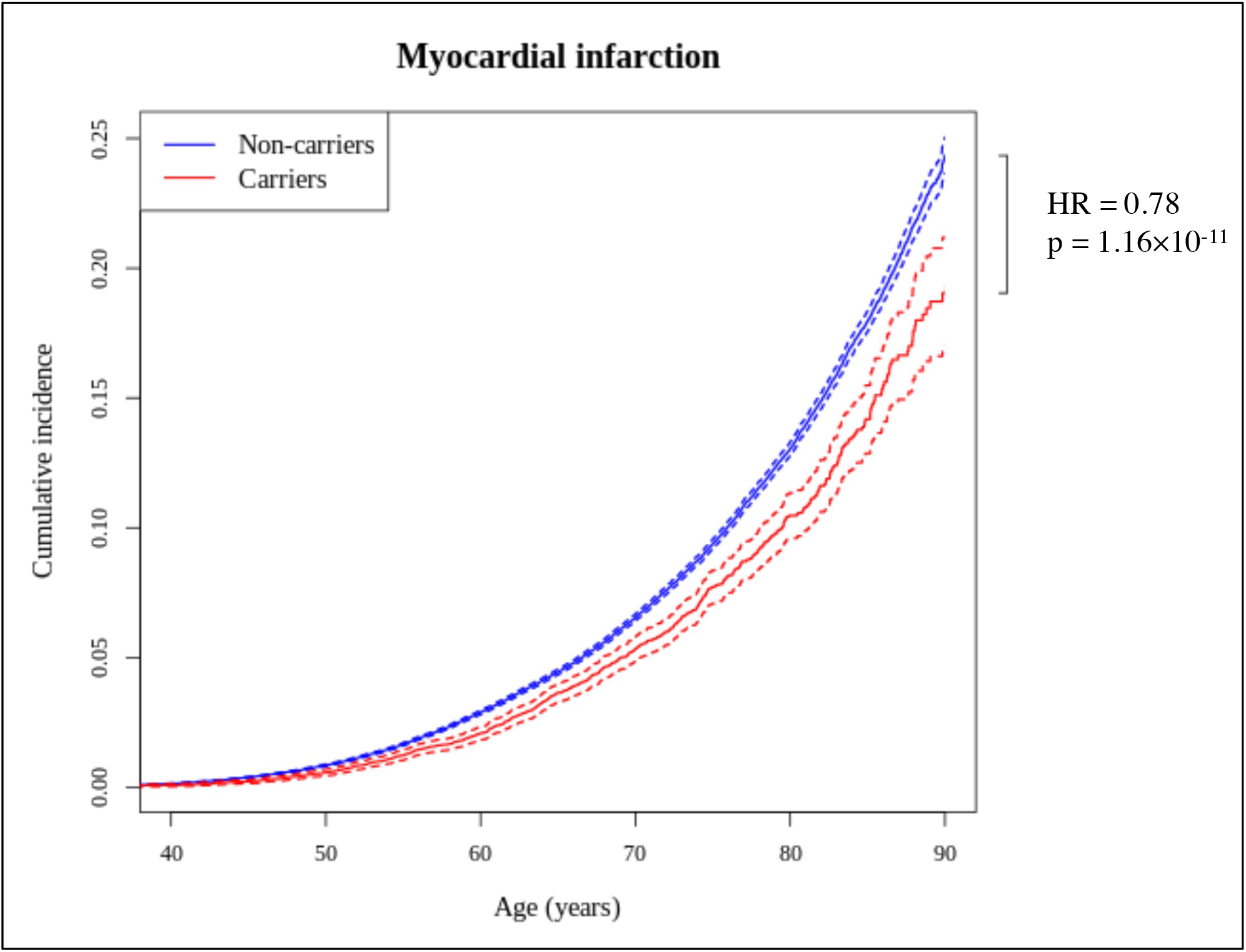
Cumulative incidence plots for myocardial infarction. Red line represents carriers (homo- or heterozygous) for either rs534125149 or rs201988637, and blue line represent non-carriers.

Dashed lines represent 95%- confidence intervals.

### Associations with risk factors for CVD and coronary heart disease sub-types

We then tested for possible associations between the *MFGE8* variants and risk factors for CVD. The splice acceptor variant rs201988637 was associated with pulse pressure in analyses across four cohorts with pulse pressure measurements and variant rs201988637 available, with the risk lowering allele associated with lower pulse pressure (p = 1.08×10^-03^, β = -1.52 [-2.43- -0.61]) mmHg (**Figure 5**). In addition, rs534125149 was significantly associated with height, but further analysis pointed this signal to be reflecting the association of a known association of *ACAN* with height, located near *MFGE8* (Supplementary material, **Supplementary Figure 8**). No associations with other risk factors were observed.

**Figure 5:**
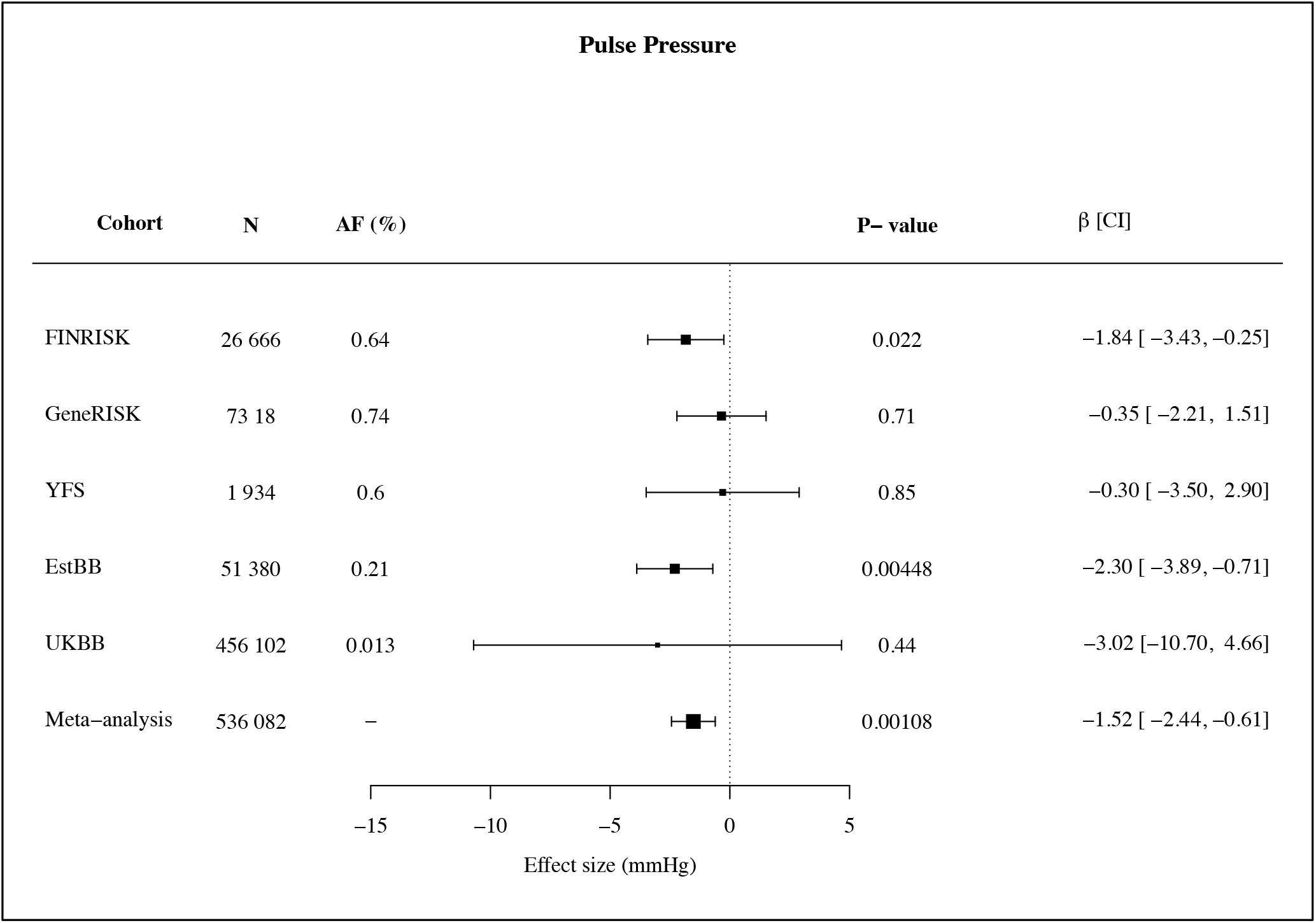
Results for pulse pressure association across all cohorts with splice acceptor variant rs201988637 present.

In the Corogene cohort (n = 4 896), rs534125149 significantly (p < 0.05) lowers the risk for acute coronary syndrome and stable coronary heart disease (RR = 0.87 and 0.83, respectively) compared to healthy controls, but not against other types of heart attack, further pointing to a specific risk- lowering effect of *MFGE8* on ischemic heart diseases (**Supplementary Figure 9**).

### Previously reported common variants near *MFGE8*

Previously, common intergenic variant (rs8042271) near *MFGE8* has been reported to associate with coronary heart disease (CHD) risk^3, 27^. We replicate this association (OR = 0.90, p = 3.69×10^-10^ for coronary atherosclerosis) in FinnGen. LD between the common variant rs8042271 and the inframe insertion rs534125149 is 0.154. The LD characteristics for all 3 variants in *MFGE8* (rs534125149, rs201988637 and rs8042271) in FinnGen are in **Supplementary Table 2**. Common variant rs8042271 was in the 95% credible set for MI with the causal probability of 0.003 but was not included in the 95% credible sets for coronary atherosclerosis (**Supplementary Table 3** and **Supplementary Table 4**). The conditional analyses of all three *MFGE8* variants showed that the association of the previously reported common variant rs8042271 can be explained by the inframe insertion variant rs534125149, but not vice versa, and that the association of the splice acceptor variant rs201988637 is independent of both rs534125149 and rs8042271 (**Supplementary Table 5**).

### Fine-mapping of the *MFGE8* locus

In our fine-mapping analyses, MI had most probably one credible set (set of causal variants) of 32 variants with the highest posterior probability (posterior probability = 0.62), and coronary atherosclerosis had two credible sets of 6 and 45 variants, respectively, with the highest posterior probability (posterior probability = 0.74). For both MI and coronary atherosclerosis, rs534125149 had the highest probability of being causal (probability of being causal = 0.250 and 0.318, respectively) and was included in the first credible set (Supplementary Table 3, **Supplementary Table 4** and **Supplementary Figure 10**). Splice acceptor variant rs201988637 was not included in the credible sets for either MI or coronary atherosclerosis, whereas previously reported common variant rs8042271 was included in the credible set for MI with the probability of being causal = 0.003 (**Supplementary Table 3**).

## Discussion

Here we show that a loss of function for *MFGE8* leads to 28% lower risk of myocardial infarction and reduces risk for other coronary atherosclerosis-related diseases. The effects are specific to coronary heart disease events and no significant association was observed to other diseases in a phenome-wide search, even if this can be due to lower statistical power in rare disease endpoints. Loss-of-function of *MFGE8* was also associated with lower pulse pressure, but not with blood lipids, blood pressure or other known coronary heart disease risk factors. In addition to *MFGE8*, we report three other novel loci associated with coronary atherosclerosis risk, and to our knowledge we report the first CHD associated locus, *FHL1,* in chromosome X.

Our findings allow us to draw several conclusions. First, *MFGE8* is a potential intervention target with specific effects on coronary heart disease. Specific protective effect with the LoF in *MFGE8* shows potential for efficacy of a treatment targeting MFGE8 protein or downstream products. Second, the lack of risk elevation in other diseases provide evidence on the potential safety of the intervention. Previously, the protective effect of loss-of-function variants have been reported for example for *PCSK9*^5^ and *APOC3*^6^, and in phase I, II and III trials, inhibition of PCSK9 have led to significantly decreased LDL-C levels, and in short-term trials, PCSK9 inhibitors have been well-tolerated and have had a low incidence of adverse effects.^28^ Based on the phenome-wide association profile for the loss-of-function variant, we hypothesize that inhibiting *MFGE8* could lower the CHD risk.

An association of loss-of-function of *MFGE8* with lower pulse pressure, a potential biomarker for arterial stiffness ^29^, are very much in line with previous studies on MFGE8 and the inflammatory ageing process of the arteries, highlighting the possible role of *MFGE8* in arterial ageing and stiffness. The *MFGE8* gene encodes Milk-fat globule-EGF 8 (MFGE8), or lactadherin, which is an integrin-binding glycoprotein implicated in vascular smooth muscle cell (VSMC) proliferation and invasion, and the secretion of pro-inflammatory molecules ^30, 31^. Lactadherin is known to play important roles in several other biological processes, including apoptotic cell clearance and adaptive immunity ^32^, which are known to contribute to the pathogenesis of ischemic stroke. Initially lactadherin was identified as a bridging molecule between apoptotic cells and phagocytic macrophages ^33–35^, but a growing evidence has indicated that it is a secreted inflammatory mediator that orchestrates diverse cellular interactions involved in the pathogenesis of various diseases, including vascular metabolic disorders and some tumors ^36–40^ and cancers, such as breast ^38, 41^, bladder^42^, esophageal^43^ and colorectal cancer ^44^. Recently, not only has MFG-E8 expression emerged as a molecular hallmark of adverse cardiovascular remodeling with age ^45–48^, but MFG-E8 signaling has also been found to mediate the vascular outcomes of cellular and matrix responses to the hostile stresses associated with hypertension, diabetes, and atherosclerosis ^49–53^.

Arterial inflammation and remodeling are linked to the pathogenesis of age-associated arterial diseases, such as atherosclerosis. Recently, lactadherin has been identified as a novel local biomarker for ageing arterial walls by high-throughput proteomic screening, and it has been shown to also be an element of the arterial inflammatory signaling network ^54^. The transcription, translation, and signaling levels of MFG-E8 are increased in aged, atherosclerotic, hypertensive, and diabetic arterial walls in vivo, as well as activated VSMCs and a subset of macrophages in vitro. During aging, both MFG-E8 transcription and translation increase within the arterial walls and hearts of various species, including rats, humans, and monkeys^48, 55–57^, and MFG-E8 is markedly up-regulated in rat aortic walls with ageing^48^. High levels of MFG-E8 have also been detected within endothelial cells, SMC, and macrophages of atherosclerotic aortae in both mice and humans^53, 58^. Furthermore, in the advanced atherosclerotic plaques found in murine models, decreased macrophage MFG-E8 levels are associated with an inhibition of apoptotic cell engulfment, leading to the accumulation of cellular debris during the pathogenesis of atherosclerosis. Lastly, lactadherin has shown tissue protection in various models of organ injury, including suppression of inflammation and apoptosis in intestinal ischemia in mice ^59^, as well as inducing recovery from ischemia by facilitating angiogenesis ^60^.

Our study does, however, have a few limitations. First, our primary association results come from Finnish population with considerable elevation in allele frequency in *MFGE8* variants among Finns. Therefore, the replication of the association in other populations has reduced statistical power. However, there were enough carriers combined in Japanese, Estonian and UK samples to replicate robustly both the protective association with coronary artery disease and for pulse pressure. Secondly, although our data shows association with pulse pressure which has previously been linked to arterial stiffness, the direct effect of the genetic variants on arterial stiffness and arterial aging needs further evidence.

In conclusion, our results show that inhibiting production of lactadherin could reduce the risk for coronary atherosclerosis substantially and thus present *MFGE8* as a potential therapeutical target for atherosclerotic cardiovascular disease. Our study also highlights the potential of FinnGen, as a large-scale biobank study in isolated population to identify high-impact variants either very rare or absent in other populations.

## Supporting information

Complete supplementary material

## Data Availability

The FinnGen data may be accessed through Finnish Biobanks' FinnBB portal (www.finbb.fi) and THL Biobank data through THL Biobank (https://thl.fi/en/web/thl-biobank).

## Acknowledgements

We would like to thank all participants of all study cohorts for their generous participation. We also want to thank Dr. Kaoru Ito at RIKEN Center for Integrative Medical Sciences for supporting the study. This work was supported by the Academy of Finland Center of Excellence in Complex Disease Genetics (Grant No 312062 and 336820 to S.R.), the Finnish Foundation for Cardiovascular Research, the Sigrid Juselius Foundation, University of Helsinki HiLIFE Fellow, Grand Challenge grants and Horizon 2020 Research and Innovation Programme (grant number 101016775 “INTERVENE” to S.R.), Academy of Finland grant number 331671 to N.M., the European Union through the European Regional Development Fund (Project No. 2014-2020.4.01.16-0125 to L.M.), the Estonian Research Council grant PRG184 [to L.M.] and the Doctoral Programme in Population Health, University of Helsinki [to S.E.R.].

The FinnGen project is funded by two grants from Business Finland (HUS 4685/31/2016 and UH 4386/31/2016) and the following industry partners: AbbVie Inc., AstraZeneca UK Ltd, Biogen MA Inc., Celgene Corporation, Celgene International II Sàrl, Genentech Inc., Merck Sharp & Dohme Corp, Pfizer Inc., GlaxoSmithKline Intellectual Property Development Ltd., Sanofi US Services Inc., Maze Therapeutics Inc., Janssen Biotech Inc, and Novartis AG. Following biobanks are acknowledged for deliverig biobank samples to FinnGen: Auria Biobank (www.auria.fi/biopankki), THL Biobank (www.thl.fi/biobank), Helsinki Biobank (www.helsinginbiopankki.fi), Biobank Borealis of Northern Finland (https://www.ppshp.fi/Tutkimus-ja-opetus/Biopankki/Pages/Biobank-Borealis-briefly-in-English.aspx), Finnish Clinical Biobank Tampere (www.tays.fi/en-US/Research_and_development/Finnish_Clinical_Biobank_Tampere), Biobank of Eastern Finland (www.ita-suomenbiopankki.fi/en), Central Finland Biobank (www.ksshp.fi/fi-FI/Potilaalle/Biopankki), Finnish Red Cross Blood Service Biobank (www.veripalvelu.fi/verenluovutus/biopankkitoiminta) and Terveystalo Biobank (www.terveystalo.com/fi/Yritystietoa/Terveystalo-Biopankki/Biopankki/). All Finnish Biobanks are members of BBMRI.fi infrastructure (www.bbmri.fi) and FinBB (https://finbb.fi/).

## Estonian Biobank research team

Tõnu Esko,

Andres Metspalu,

Reedik Mägi,

Mari Nelis

## Contributors of FinnGen

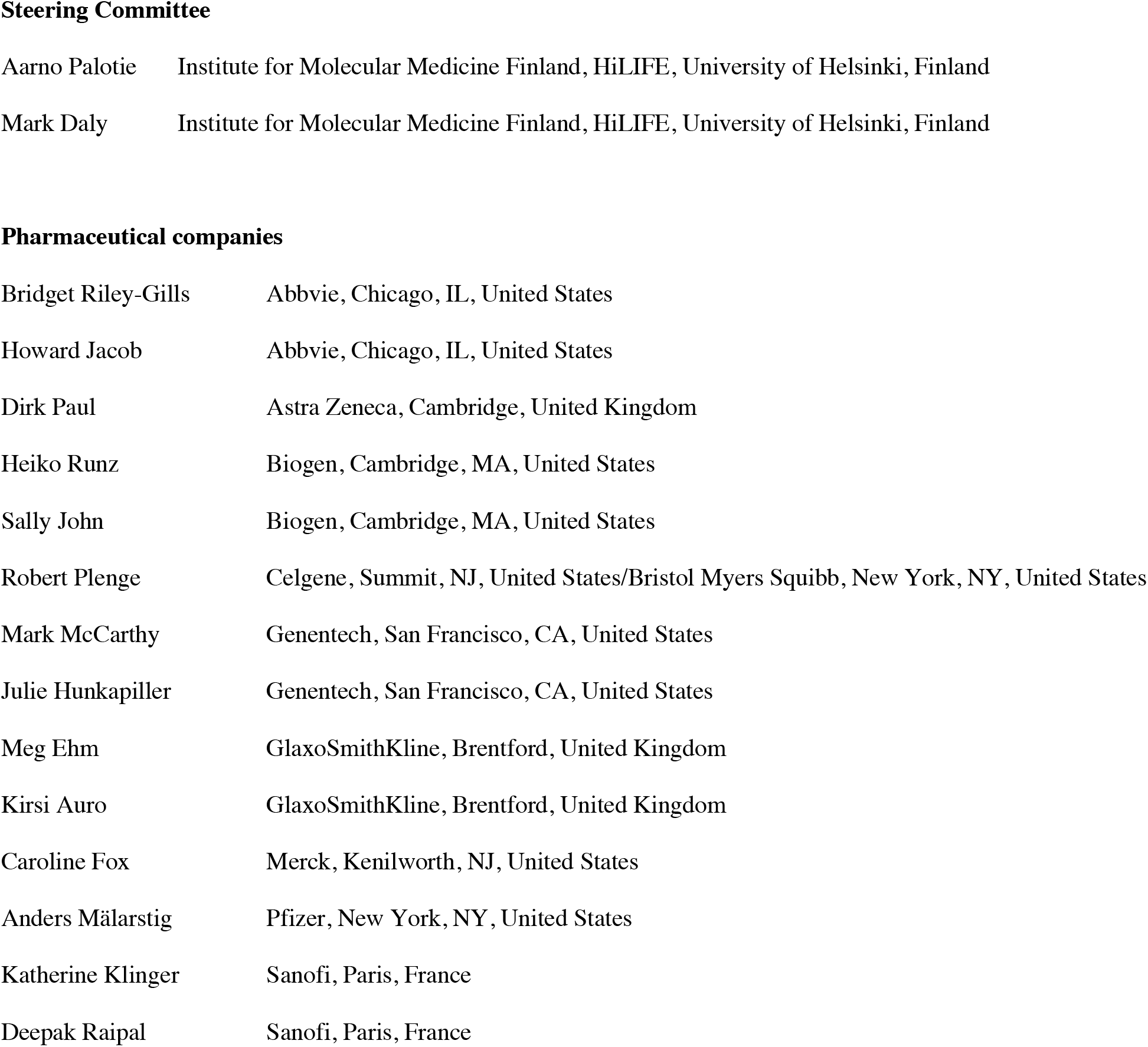
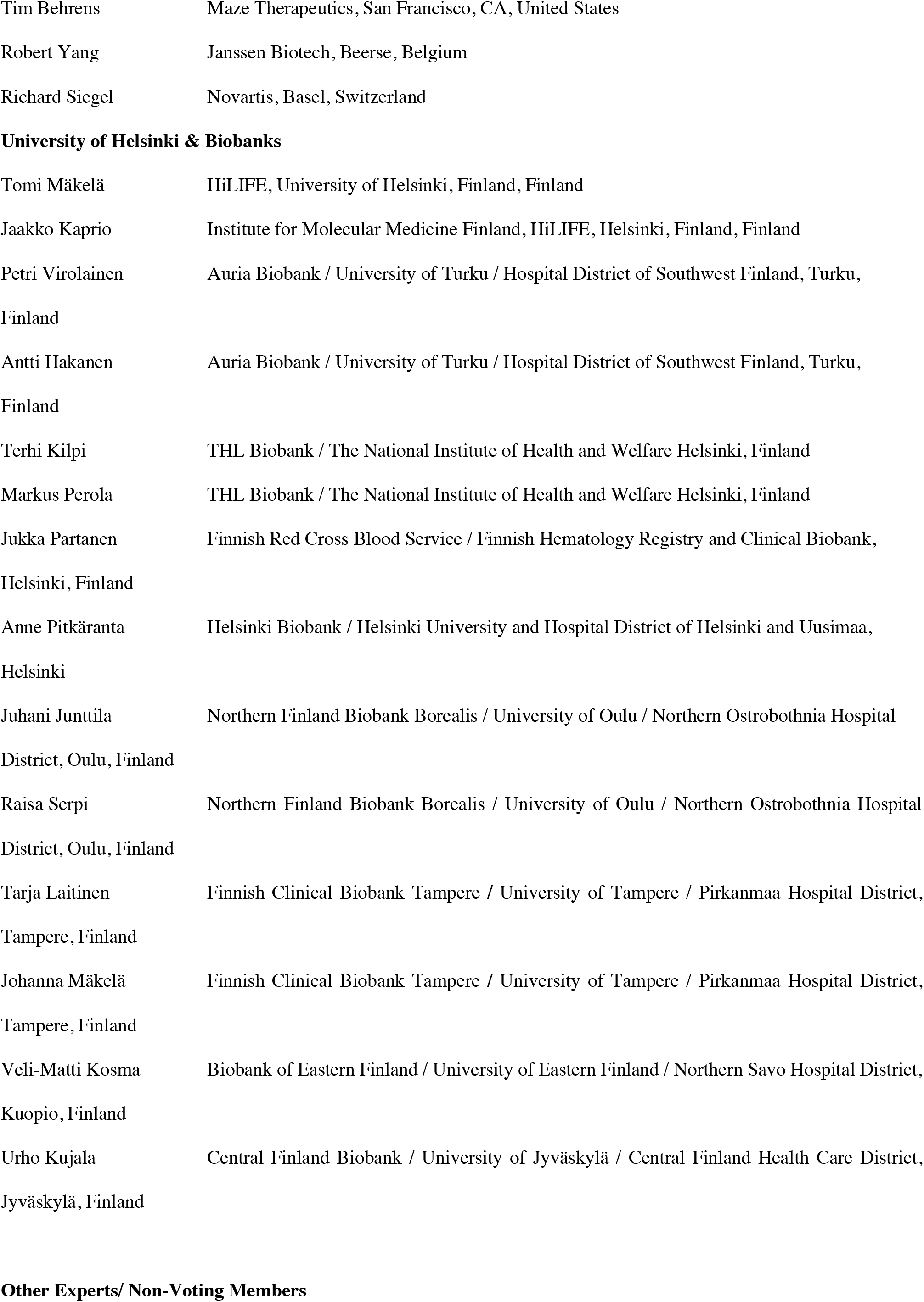
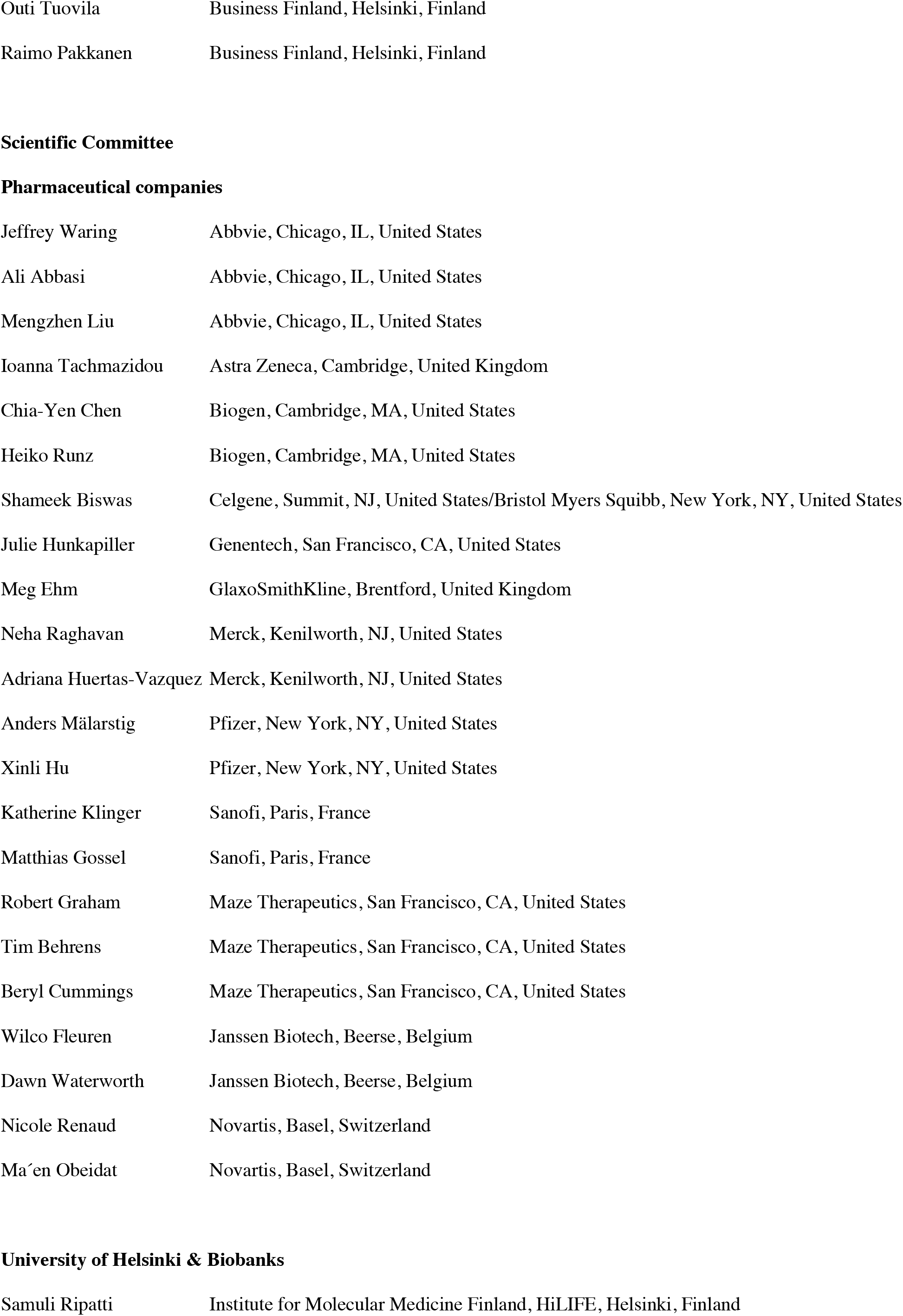
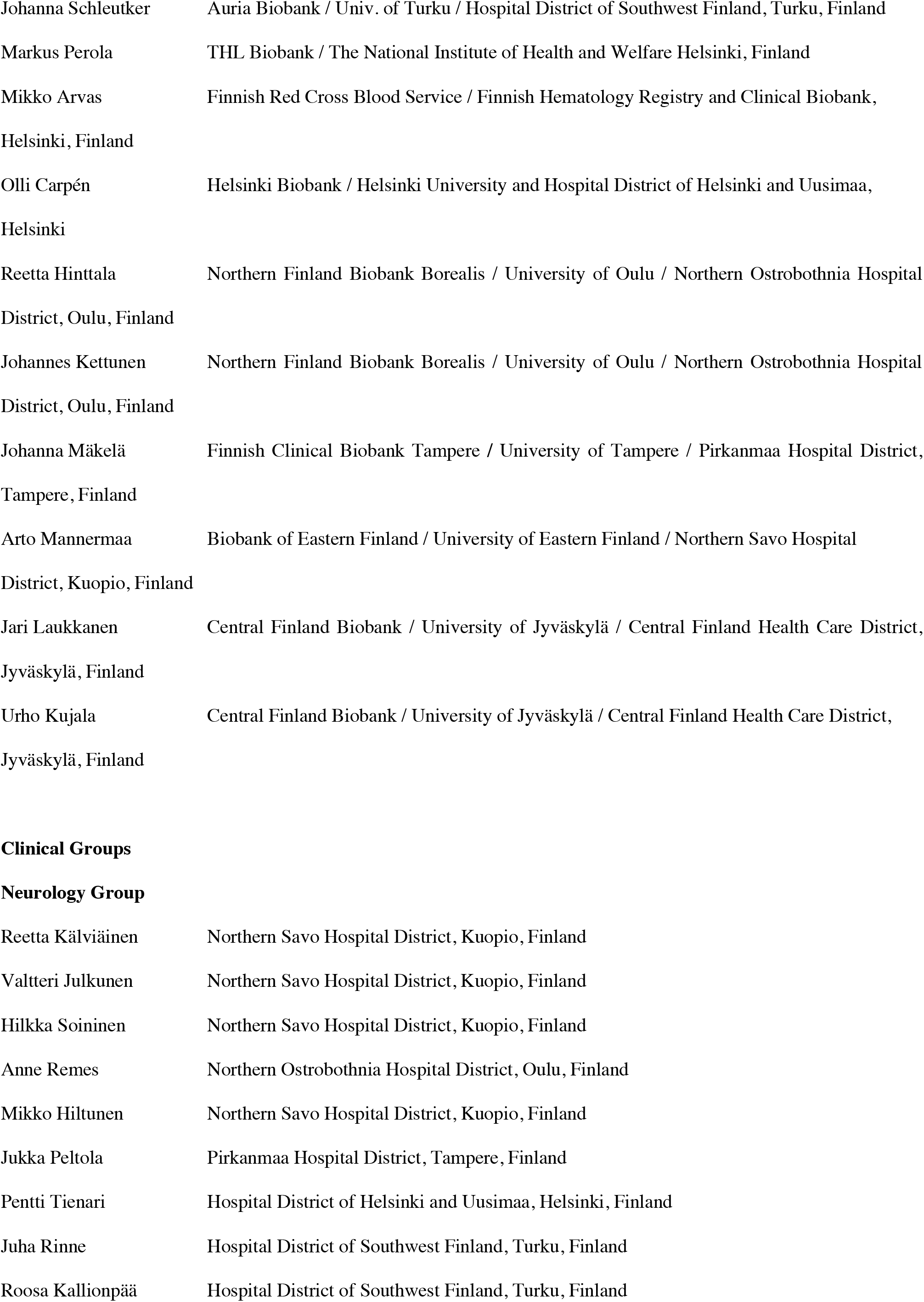
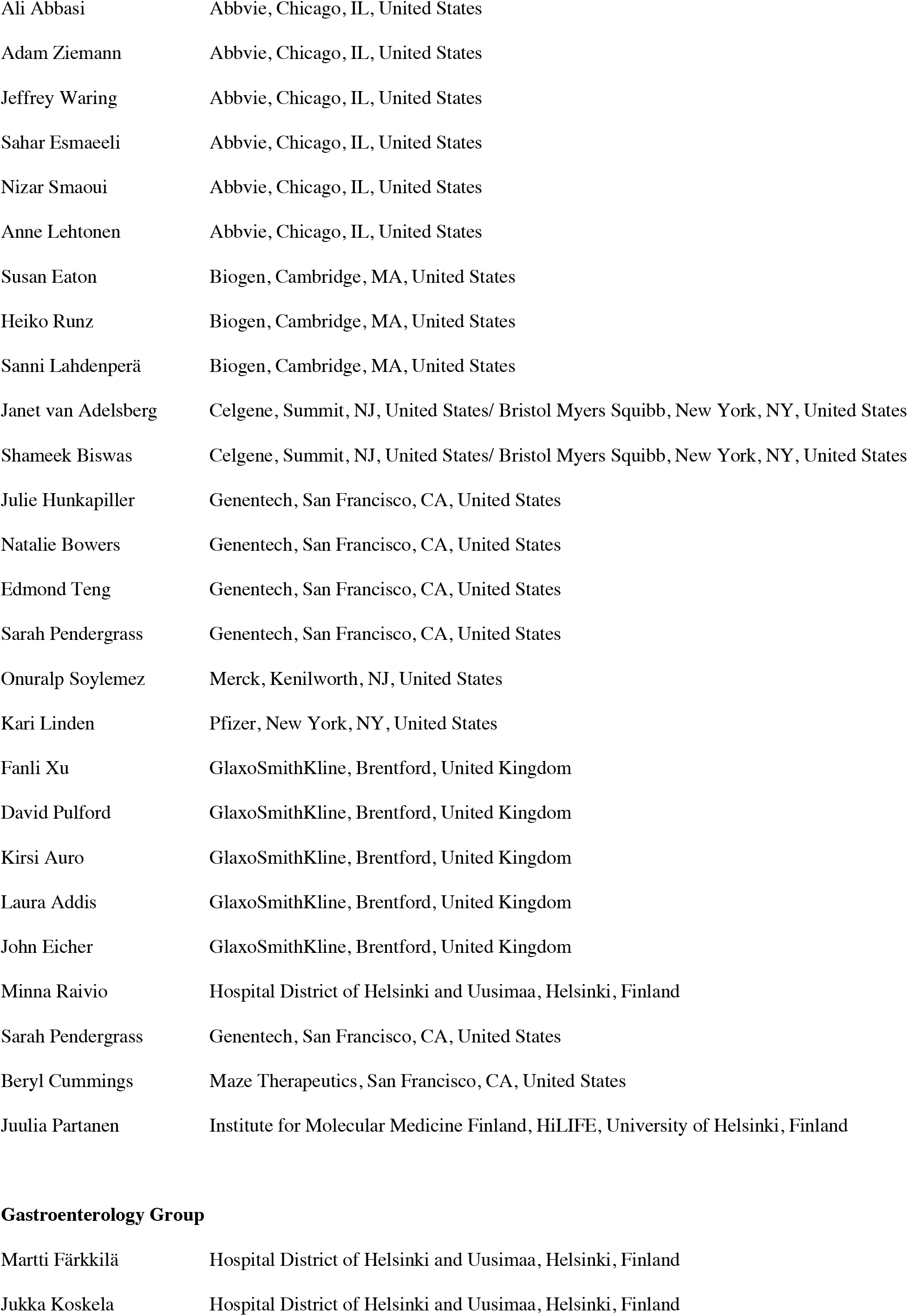
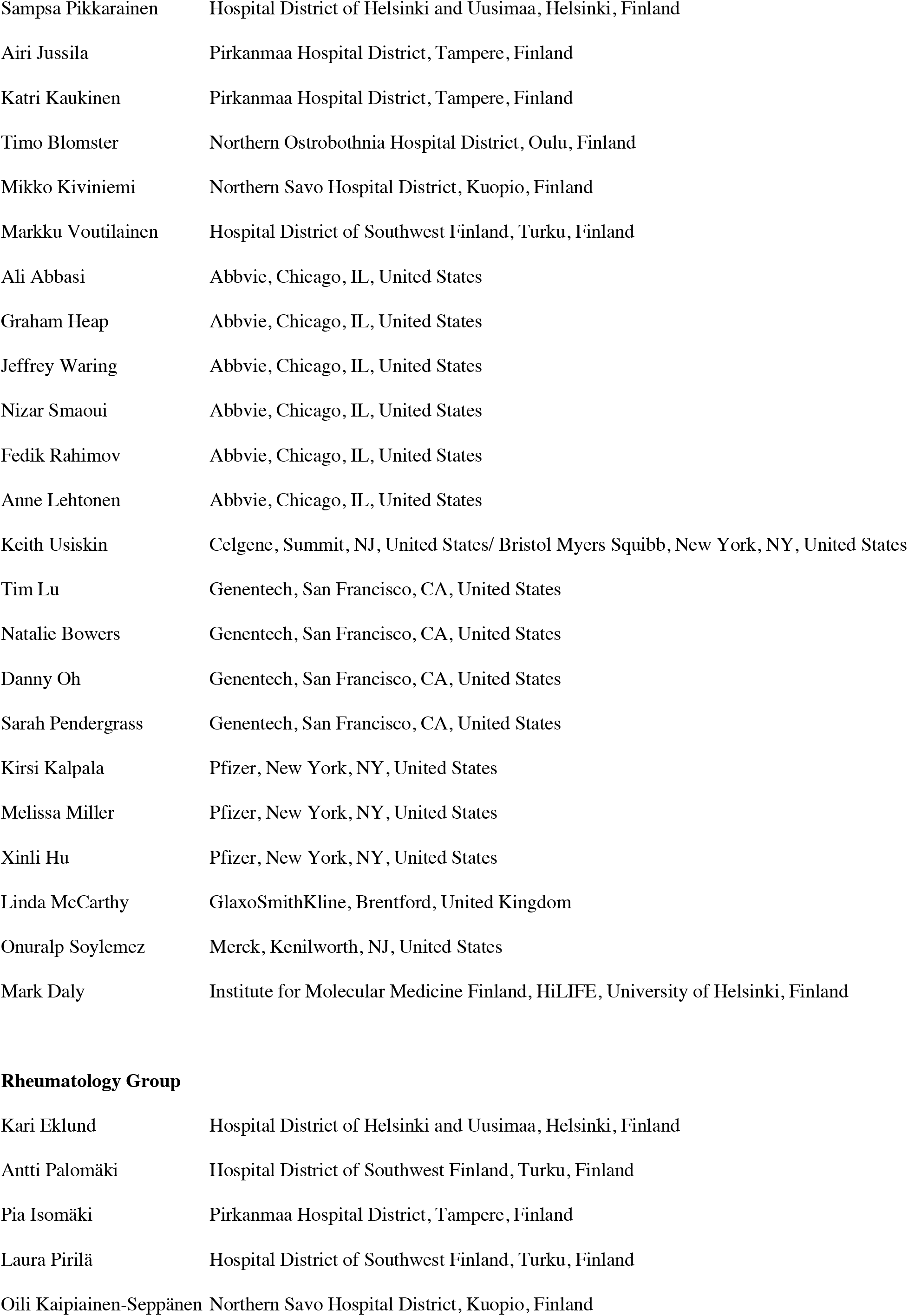
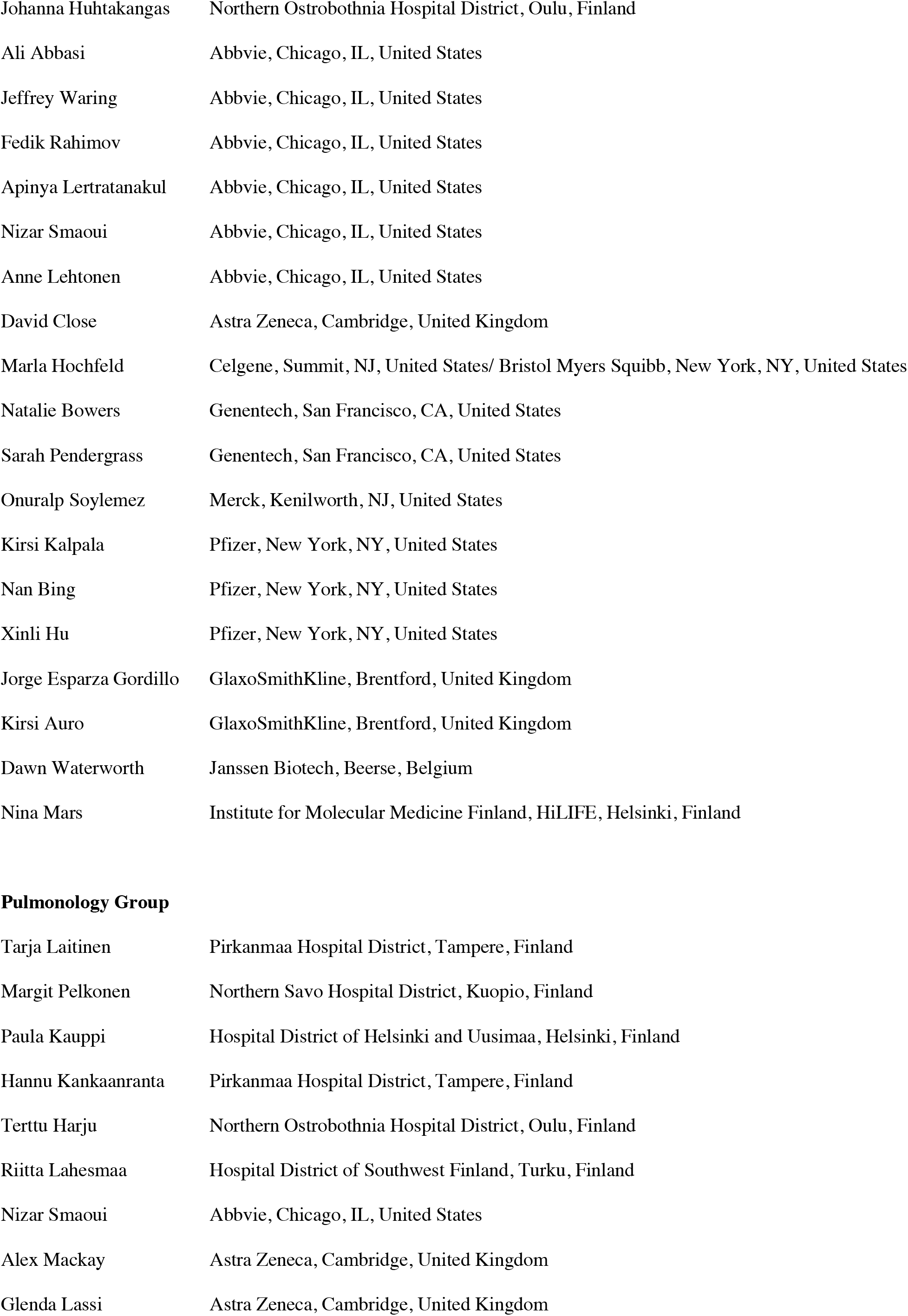
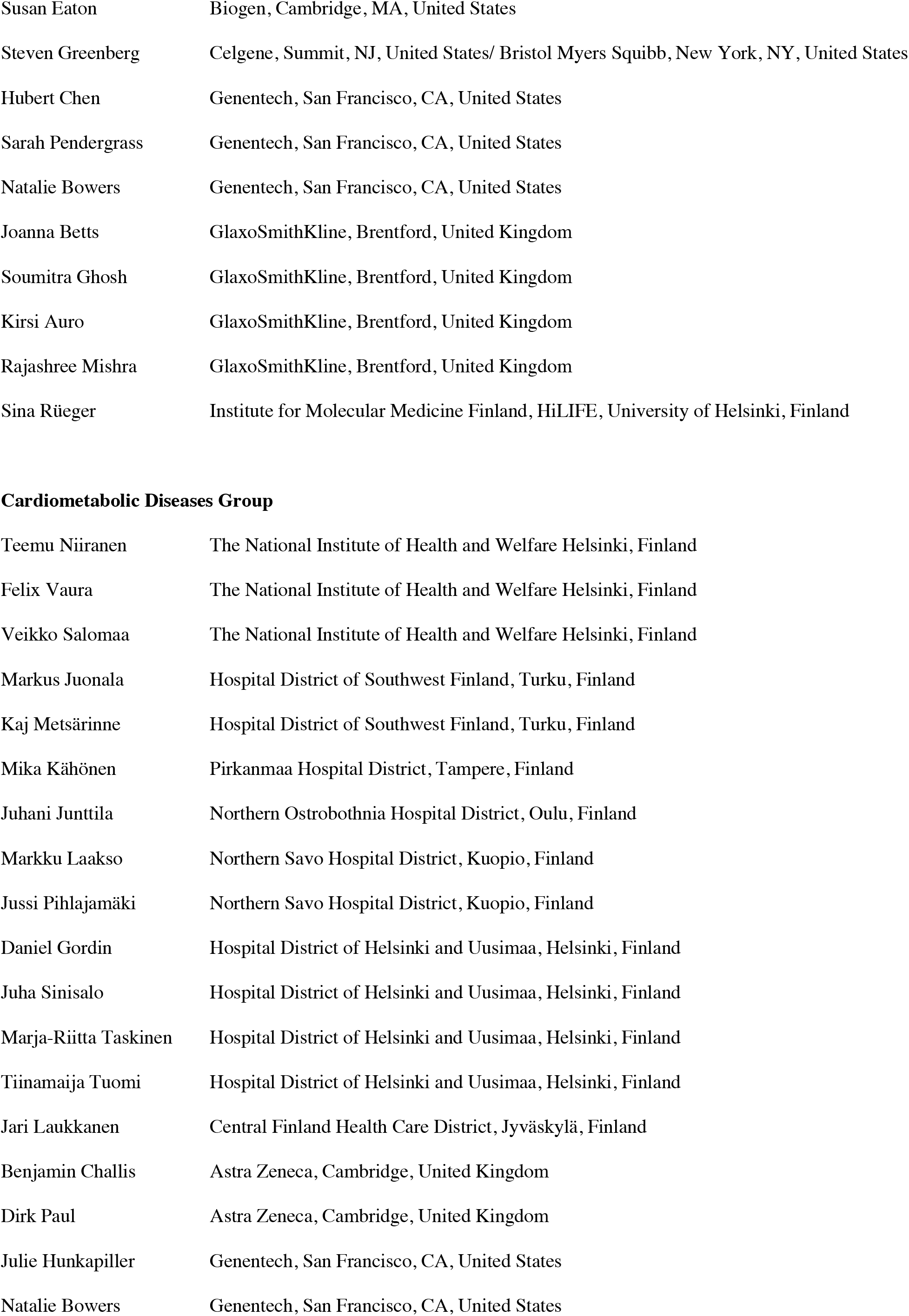
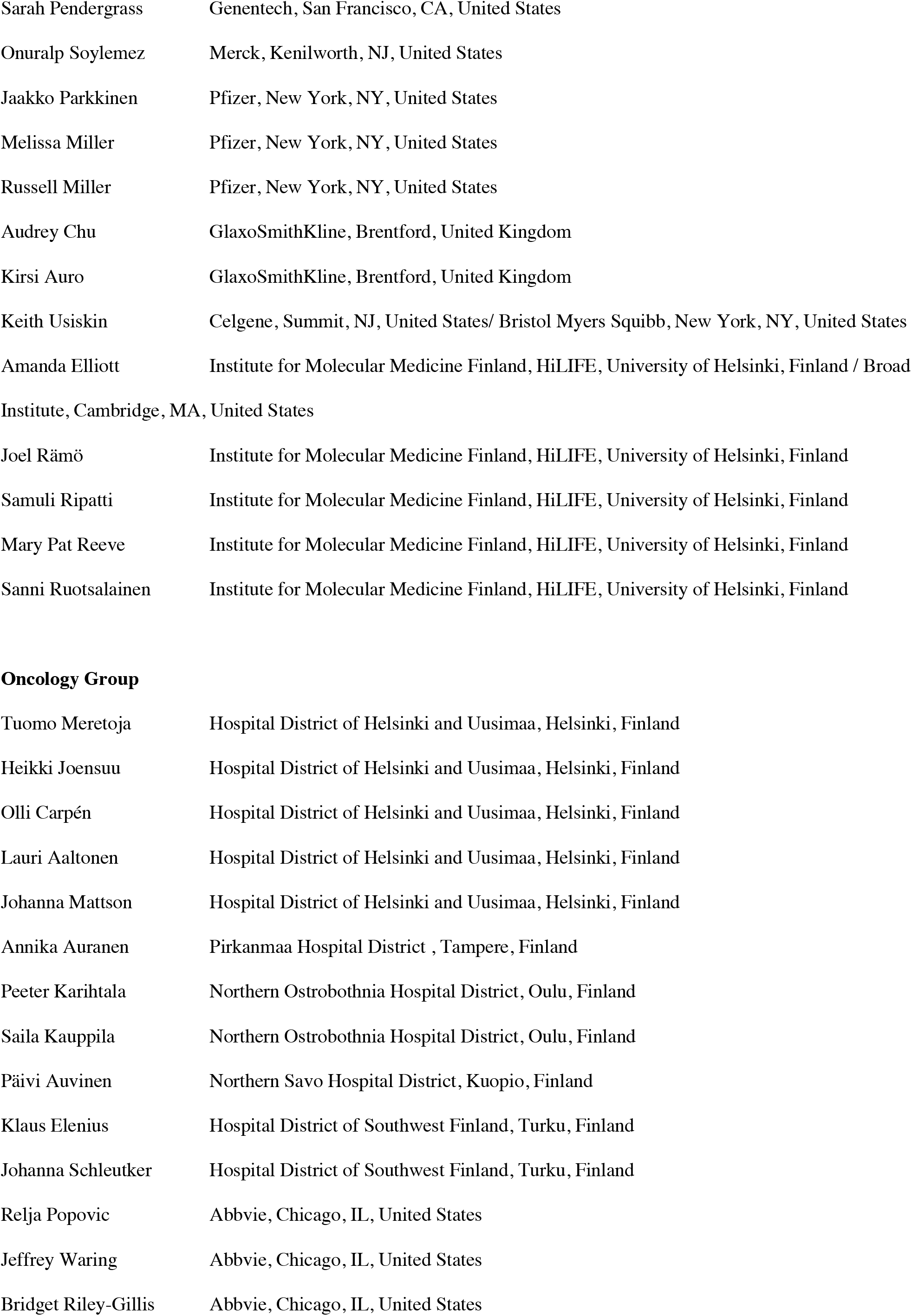
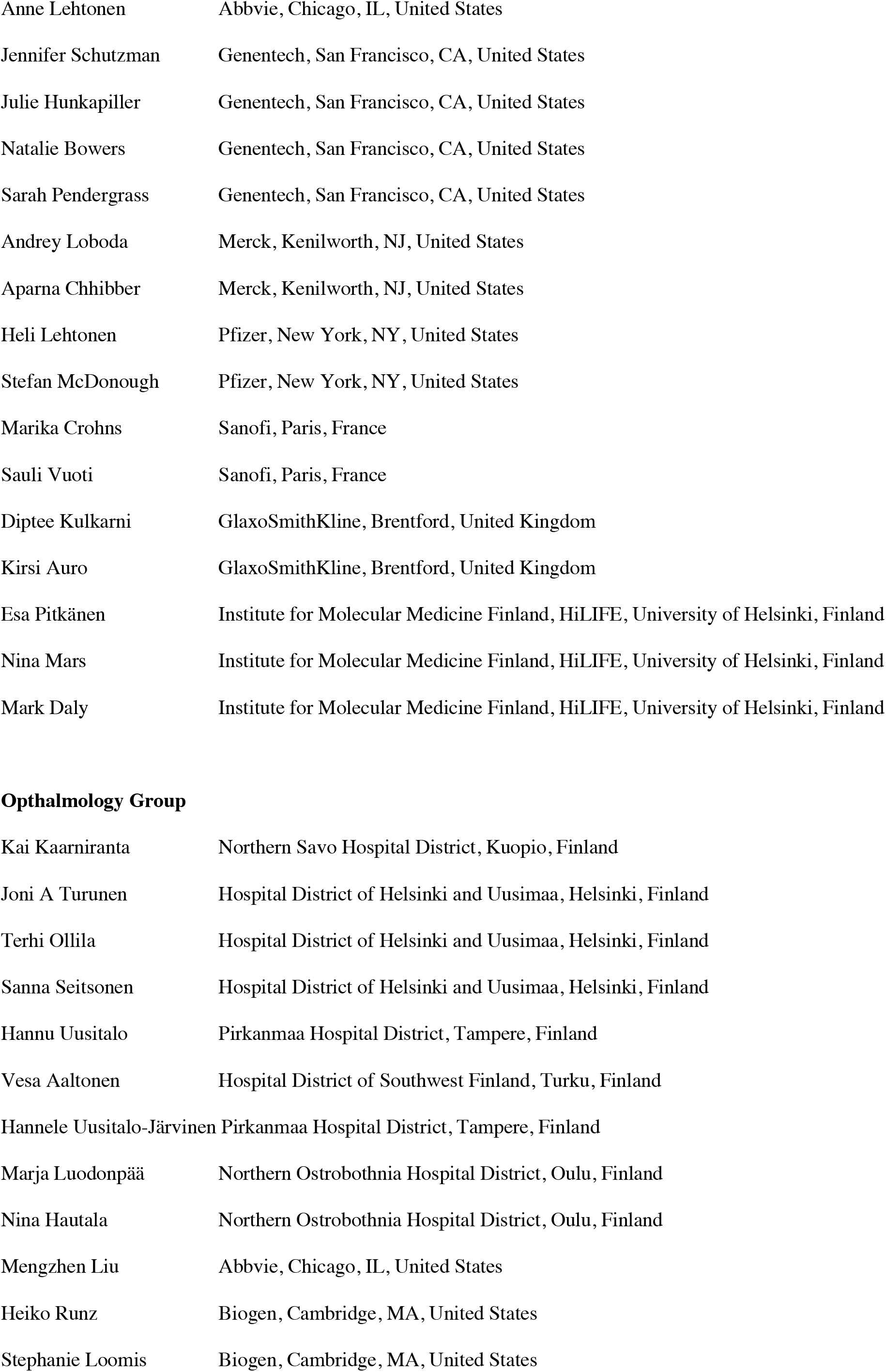
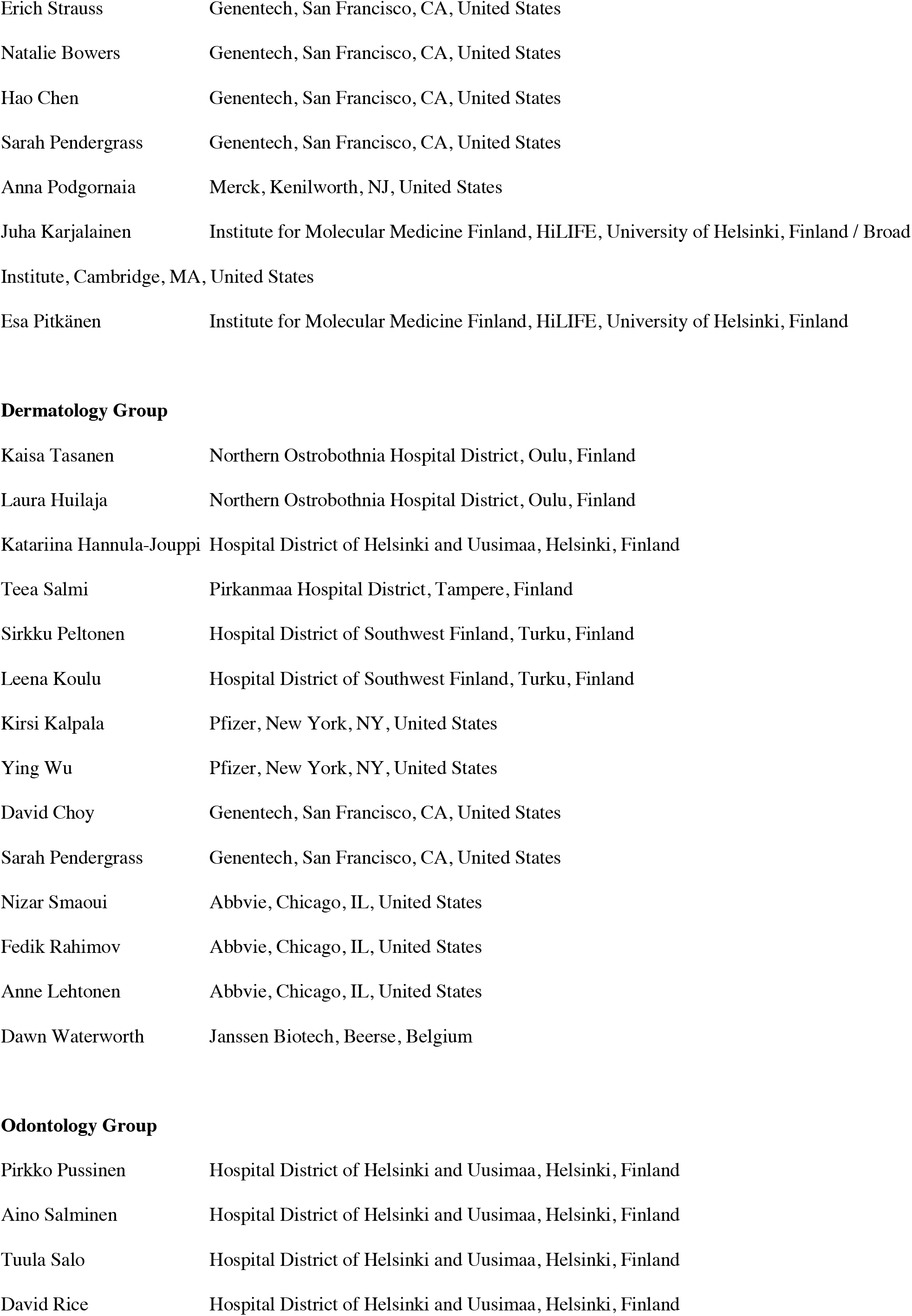
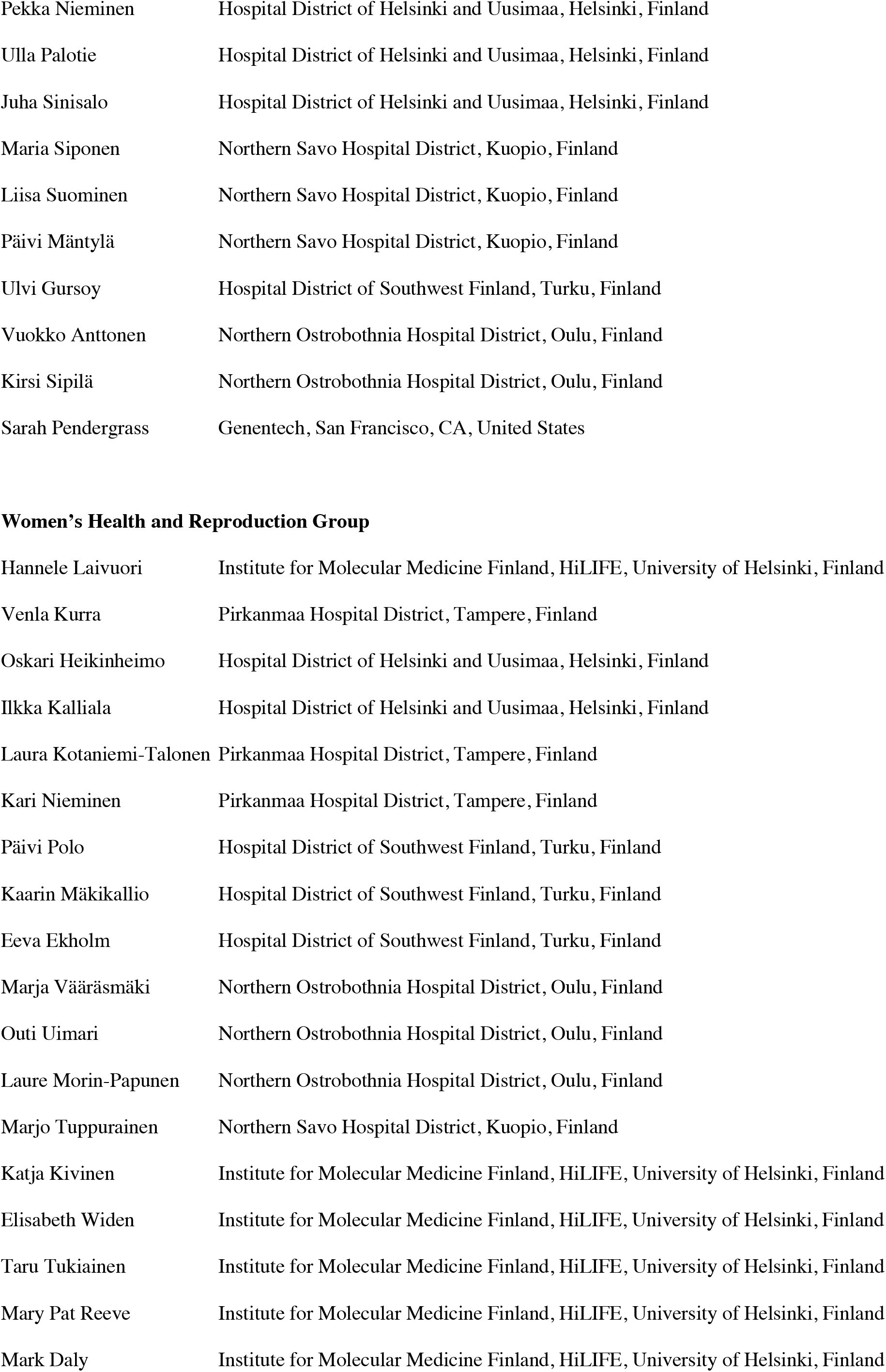
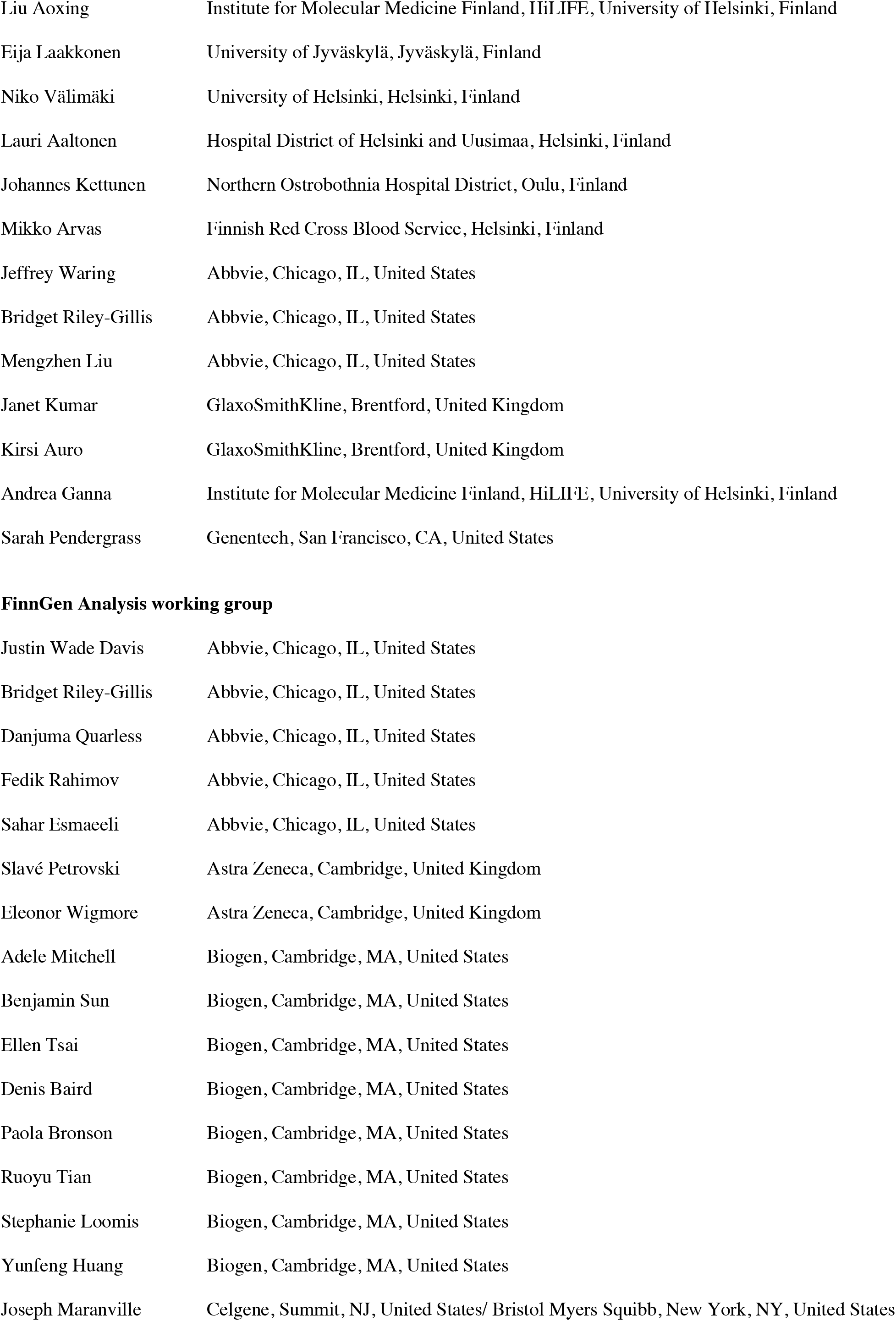
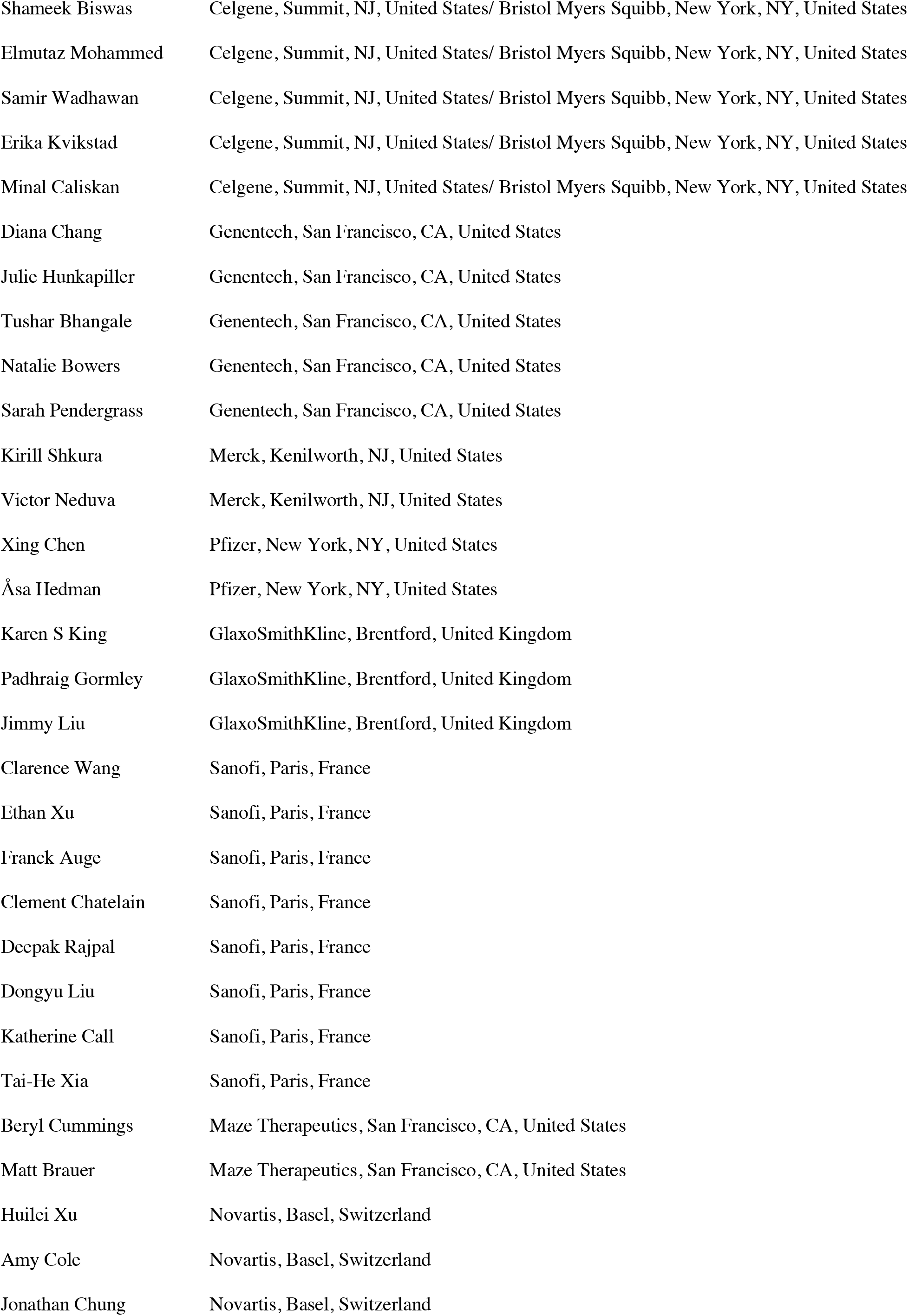
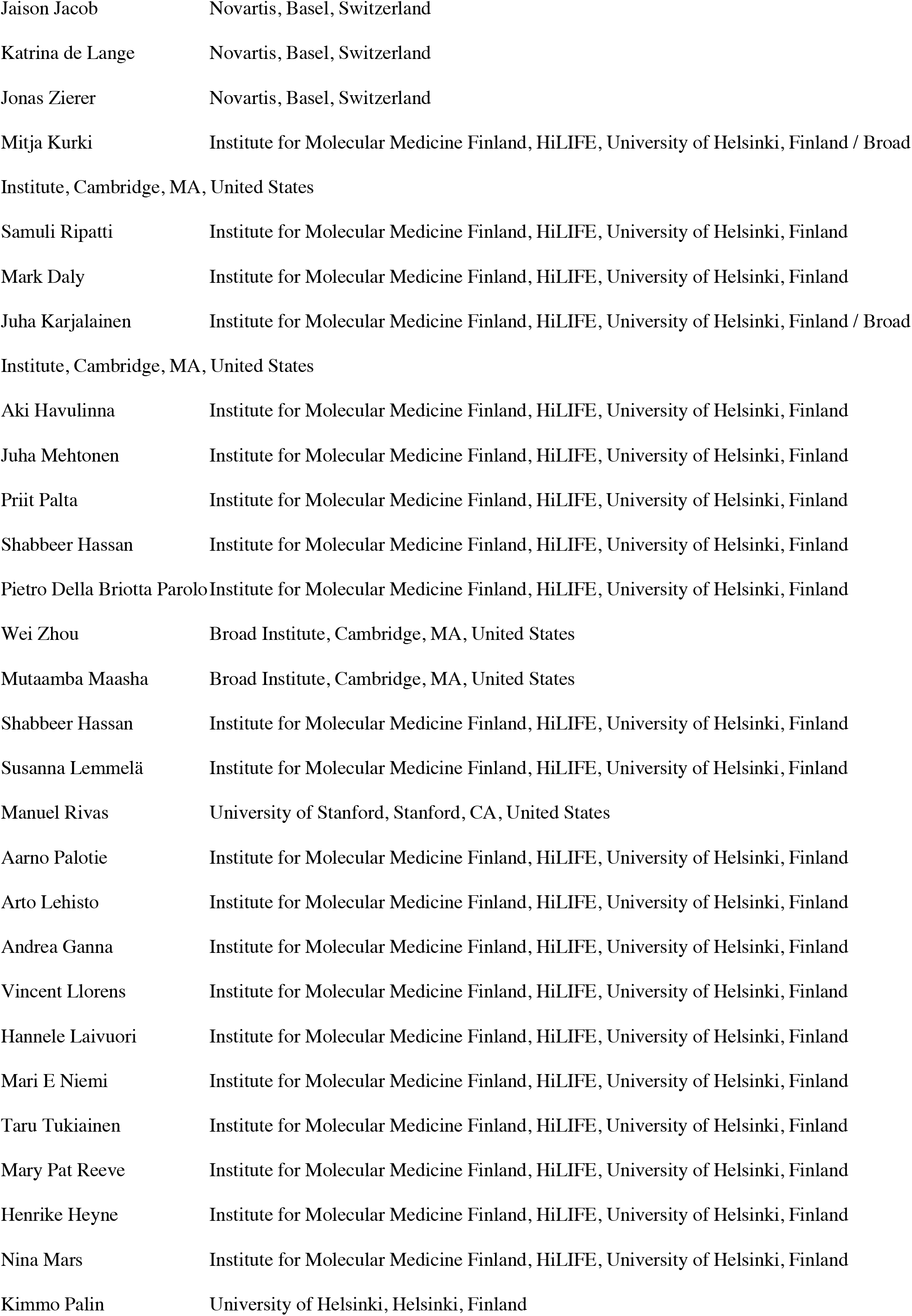
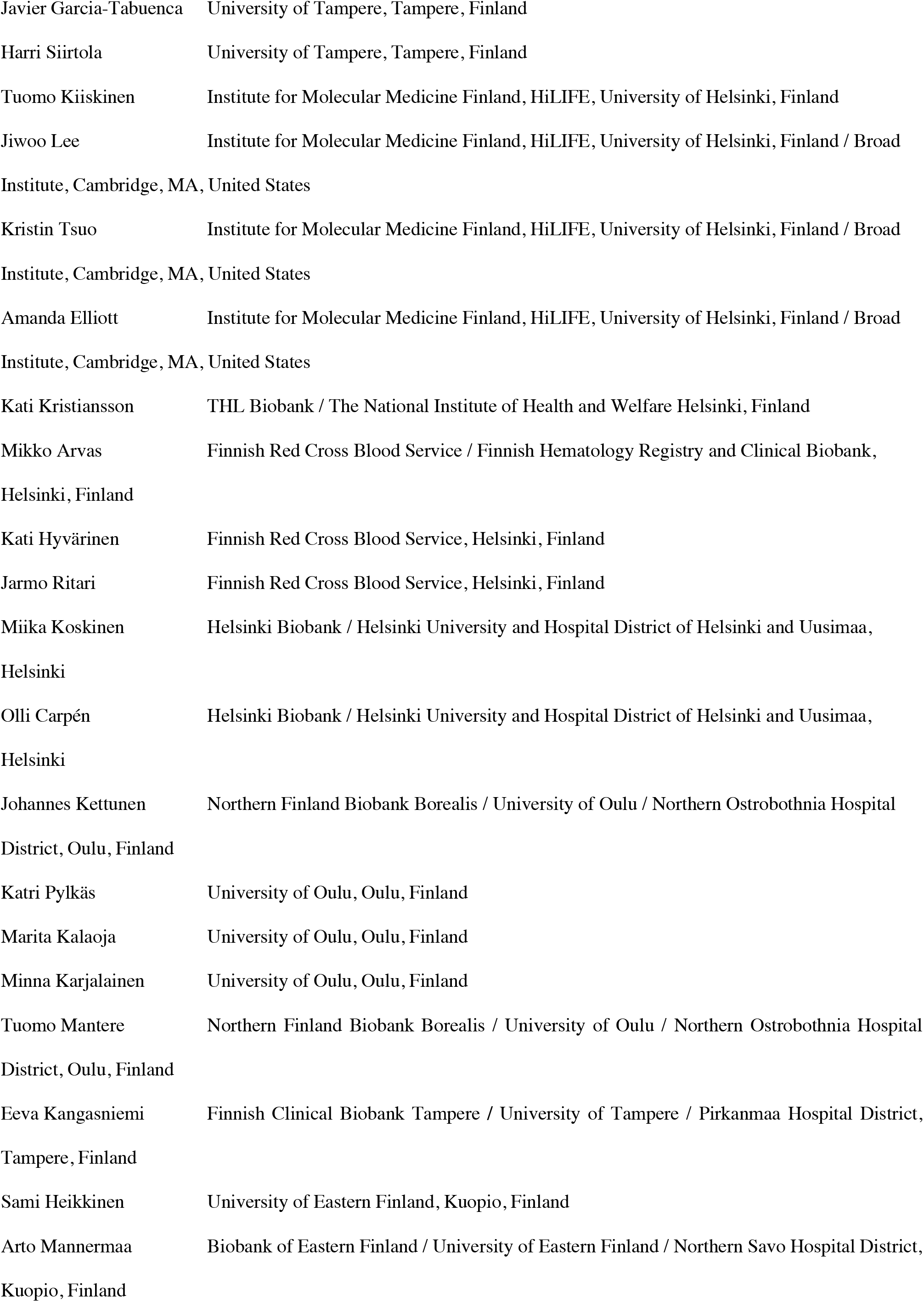
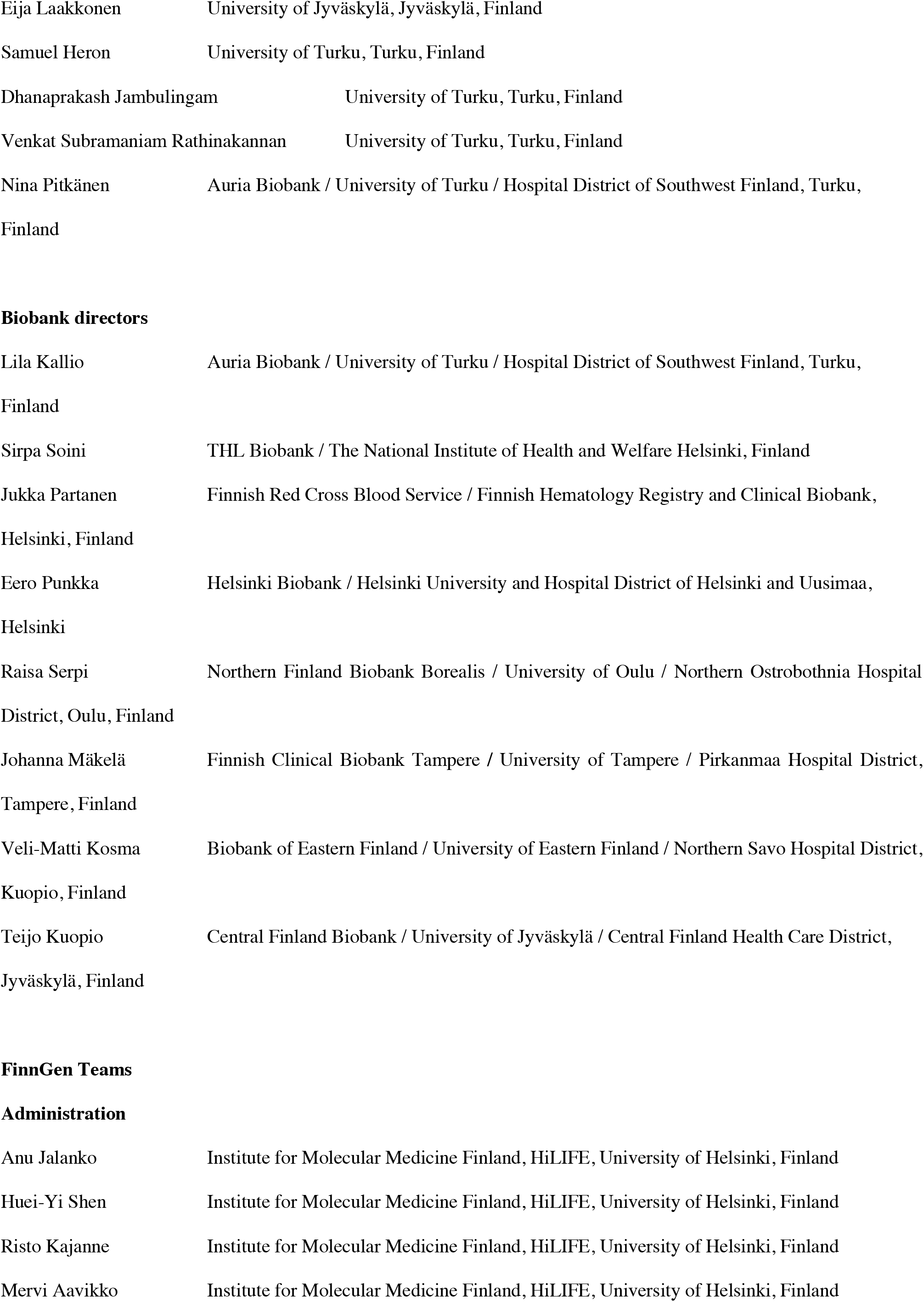
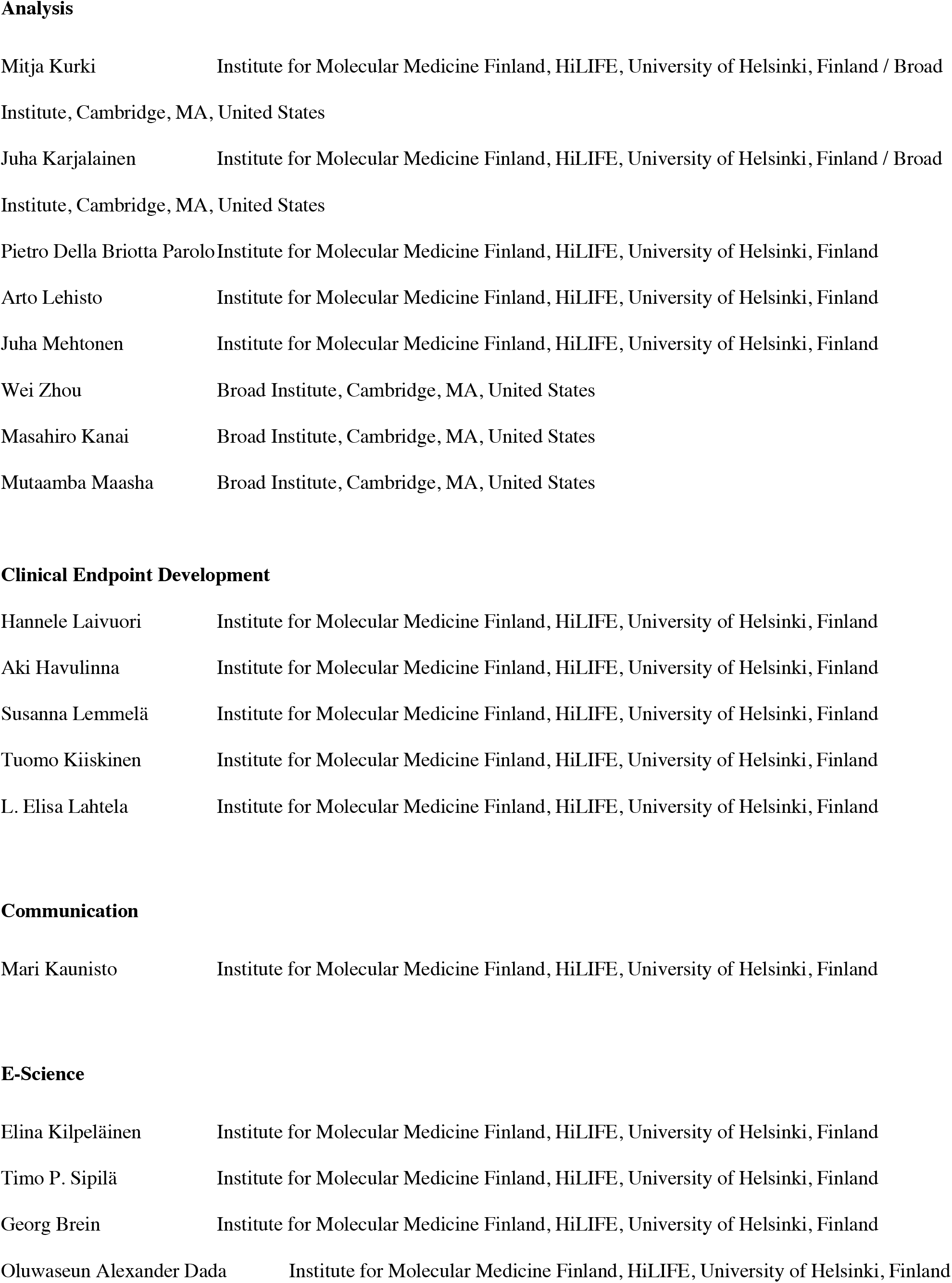
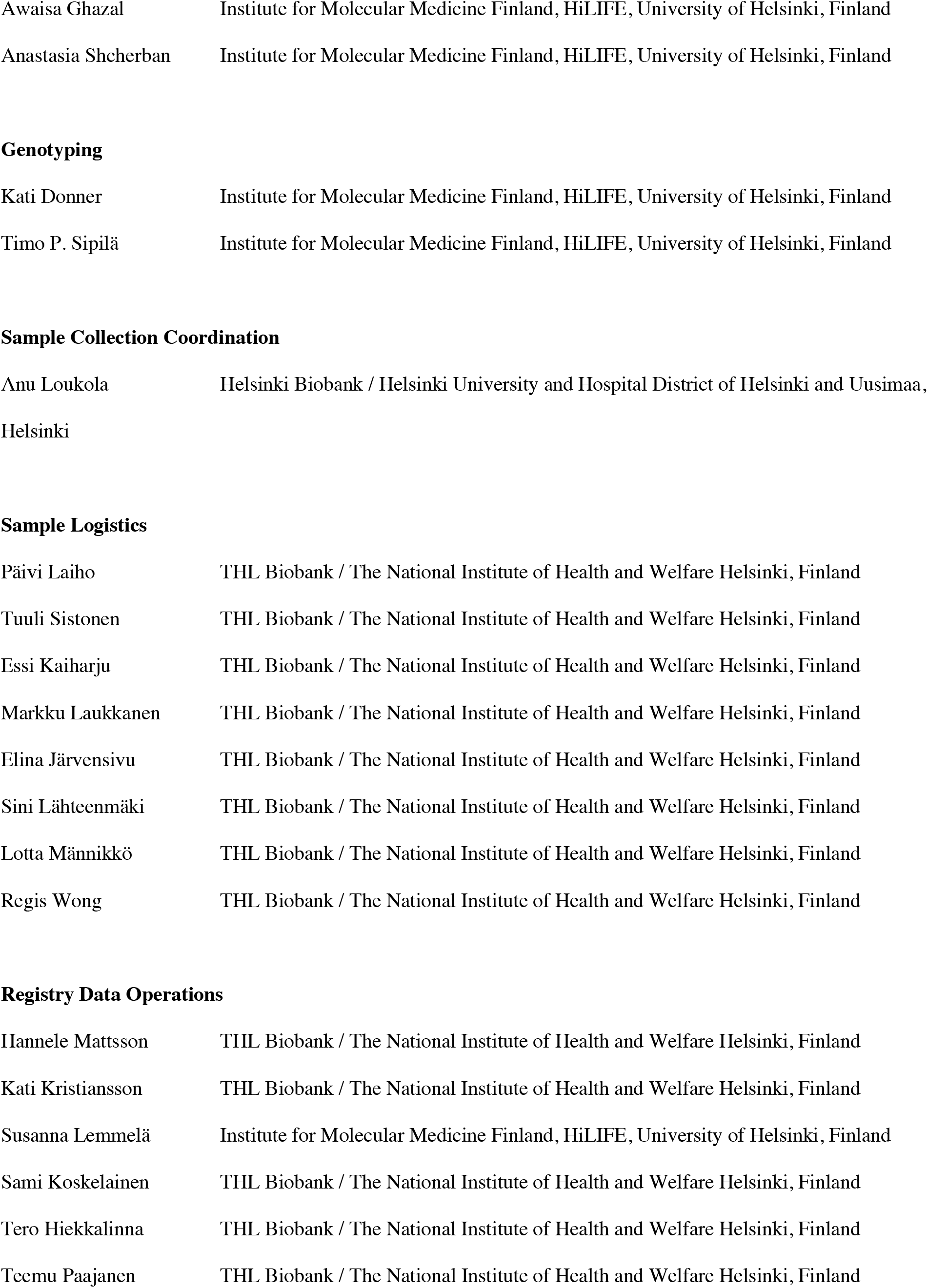
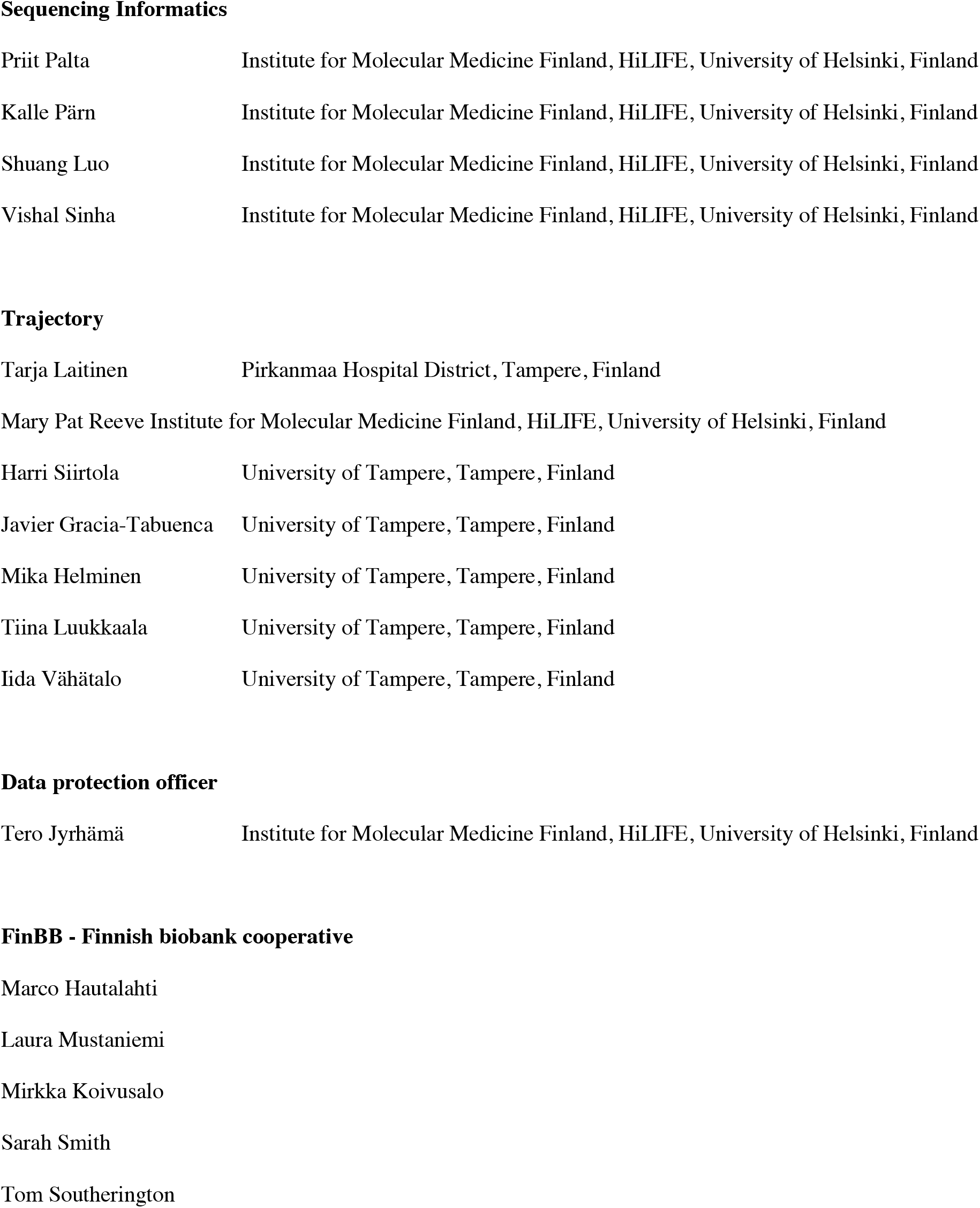

## Ethics statement

Patients and control subjects in FinnGen provided informed consent for biobank research, based on the Finnish Biobank Act. Alternatively, separate research cohorts, collected prior the Finnish Biobank Act came into effect (in September 2013) and start of FinnGen (August 2017), were collected based on study-specific consents and later transferred to the Finnish biobanks after approval by Fimea, the National Supervisory Authority for Welfare and Health. Recruitment protocols followed the biobank protocols approved by Fimea. The Coordinating Ethics Committee of the Hospital District of Helsinki and Uusimaa (HUS) approved the FinnGen study protocol Nr HUS/990/2017.

The FinnGen study is approved by Finnish Institute for Health and Welfare (permit numbers: THL/2031/6.02.00/2017, THL/1101/5.05.00/2017, THL/341/6.02.00/2018, THL/2222/6.02.00/2018, THL/283/6.02.00/2019, THL/1721/5.05.00/2019, THL/1524/5.05.00/2020, and THL/2364/14.02/2020), Digital and population data service agency (permit numbers: VRK43431/2017-3, VRK/6909/2018-3, VRK/4415/2019-3), the Social Insurance Institution (permit numbers: KELA 58/522/2017, KELA 131/522/2018, KELA 70/522/2019, KELA 98/522/2019, KELA 138/522/2019, KELA 2/522/2020, KELA 16/522/2020 and Statistics Finland (permit numbers: TK-53-1041-17 and TK-53-90-20).

The Biobank Access Decisions for FinnGen samples and data utilized in FinnGen Data Freeze 6 include: THL Biobank BB2017_55, BB2017_111, BB2018_19, BB_2018_34, BB_2018_67, BB2018_71, BB2019_7, BB2019_8, BB2019_26, BB2020_1, Finnish Red Cross Blood Service Biobank 7.12.2017, Helsinki Biobank HUS/359/2017, Auria Biobank AB17-5154, Biobank Borealis of Northern Finland_2017_1013, Biobank of Eastern Finland 1186/2018, Finnish Clinical Biobank Tampere MH0004, Central Finland Biobank 1-2017, and Terveystalo Biobank STB 2018001.

The FINRISK data used for the study were obtained from THL Biobank with application number BB2015_55.1 and UKBB data using the UK Biobank Resource with application number 22627. The study was approved by the Estonian Committee on Bioethics and Human Research (approval number 1.1-12/624).

