## Supplementary material for "Loss-of-function of *MFGE8* and protection against coronary atherosclerosis": Complete supplementary material

#### Definition of the disease endpoints

Detailed definition for the endpoints that are discussed in this study: myocardial infarction, myocardial infarction (strict definition) and coronary atherosclerosis.

**Myocardial infarction:** Myocardial infarction was defined as underlying or direct cause of death or as the main or side diagnosis at hospital discharge with ICD codes I21 and I22 (ICD-10) or 410 (ICD-9 and 8).

**Myocardial infarction (strict definition):** Myocardial infarction was defined as underlying or direct cause of death or as the main diagnosis at hospital discharge with ICD codes I21 and I22 (ICD-10) or 410 (ICD-9 and 8). Samples with ICD codes I2190, I2197, I2290 or I2297 (ICD-10) as underlying or direct cause of death or as the main diagnosis at hospital discharge were excluded from the cases.

**Coronary atherosclerosis:** Coronary atherosclerosis was defined as underlying or direct cause of death or as the main or side diagnosis at hospital discharge with ICD codes I24, I25, Z951 and T822 (ICD-10) or 414 and 9960A (ICD-9) or 414 (ICD-8). Samples with ICD code I253 (ICD-10) as underlying or direct cause of death or as the main or side diagnosis at hospital discharge were excluded from the cases.

### Height association

To assess whether the association of rs534125149 with height was due to the *MFGE8* gene, we first performed conditional analysis of height conditioning the association for rs534125149, the lead variant in FinnGen height GWAS (rs11630187) and for previously known height-associated variant in the locus, rs16942341[55], separately. Conditioning the height association on rs534125149 did not have much effect on the association of the lead variant for height (rs11630187) in the region (p- value before conditioning = $5.07\times10^{-34}$ and after conditioning = $1.19\times10^{-26}$), whereas when conditioning on the lead variant for height (rs11630187) in the region, the smallest p-value in the region was $1.39\times10^{-15}$ (for variant rs28564751). In addition, conditioning on either known height-associated variant rs16942341 or lead variant for height in FinnGen (rs11630187) did not affect on rs534125149's association with height (p-value before conditioning = $8.04\times10^{-13}$ and after conditioning = $3.14\times10^{-12}$ and $2.75\times10^{-05}$, respectively)

In addition, to assess whether the association of rs534125149 with atherosclerotic cardiovascular disease and height reflect the same signal, we performed colocalization analysis in the region ±200kB around rs53412514 using Coloc package in R. The probability for shared signal (PP4) was $9.22\times10^{-13}$, whereas probability for two independent (PP3) signals was 1, indicating two independent signals for height and coronary atherosclerosis in the locus.

### Supplementary figures

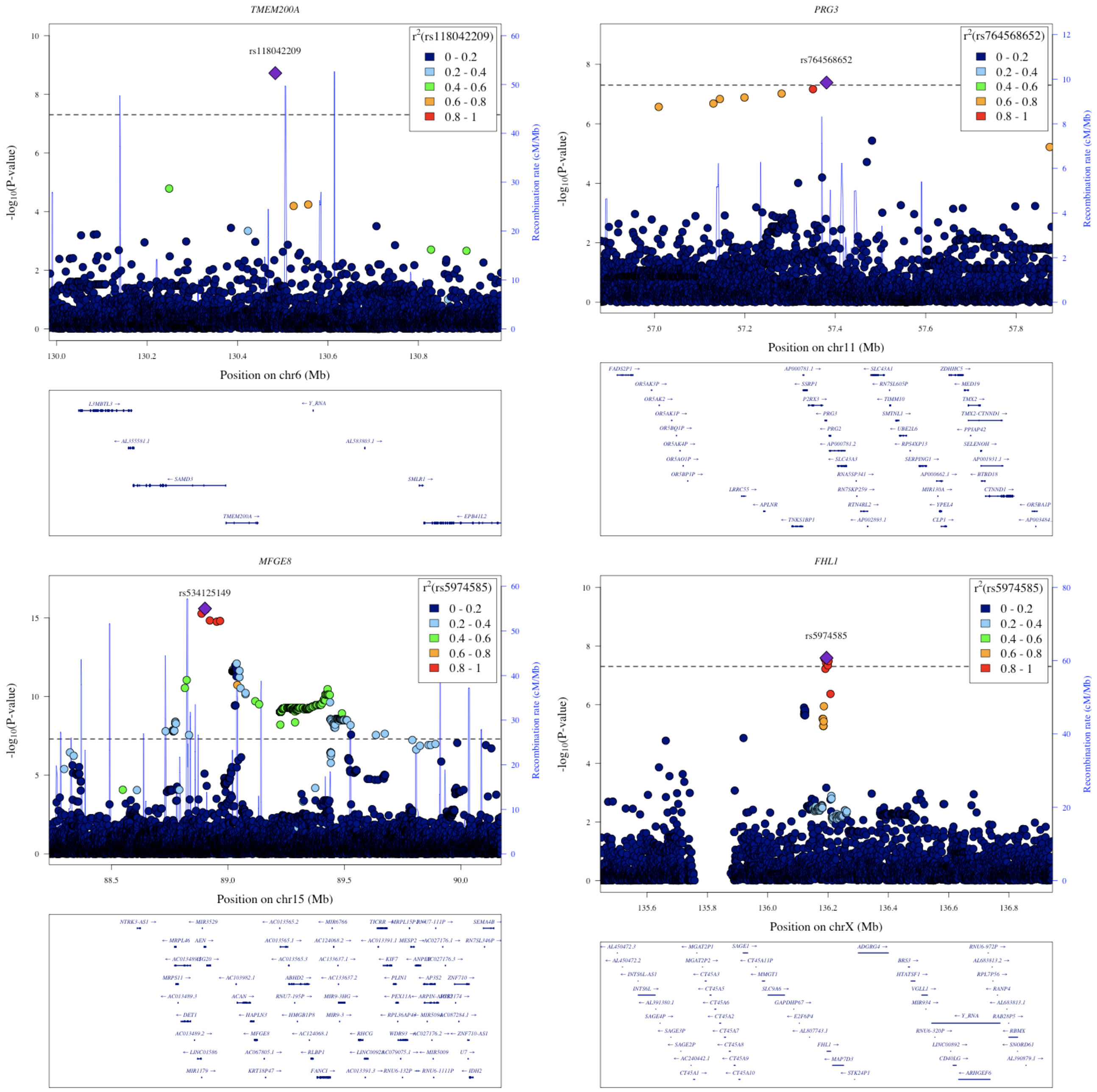


***Supplementary Figure 1:*** *Locuszoomplots for the 4 novel loci for CVD- related endpoints*.

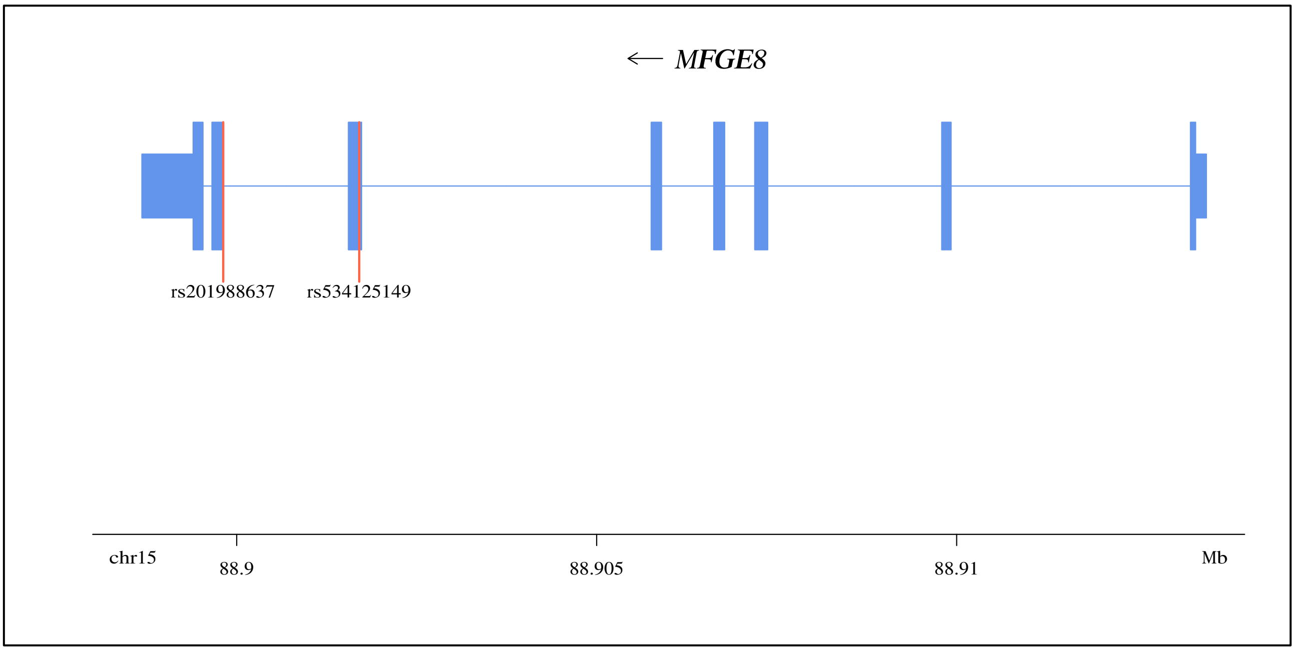


***Supplementary Figure 2:*** *MFGE8 gene structure, inframe insertion (rs534125149) and splice acceptor variant (rs201988637) highlighted.*

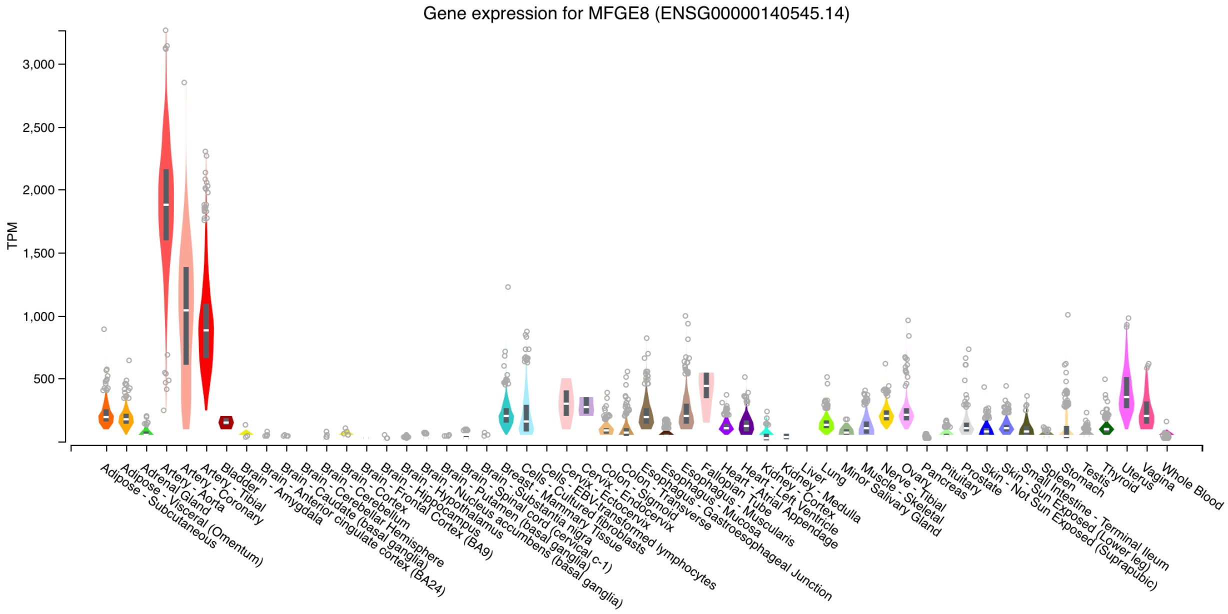


***Supplementary Figure 3:*** *Gene expression for MFGE8 gene (GTEX v8: https://gtexportal.org/home/gene/MFGE8).*

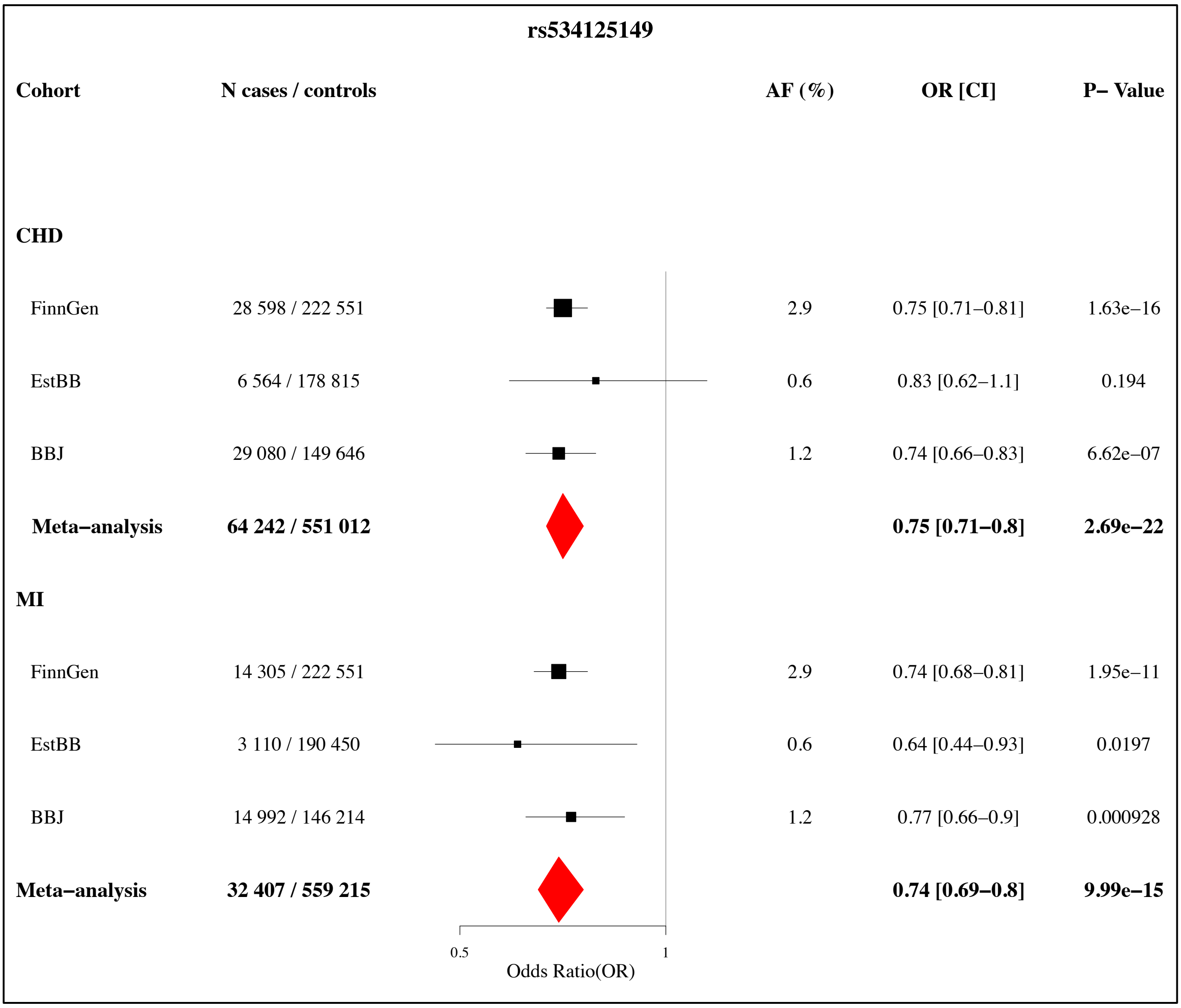


***Supplementary Figure 4:*** *Results for rs534125149 against coronary artery disease and myocardial infarction across different cohorts.*

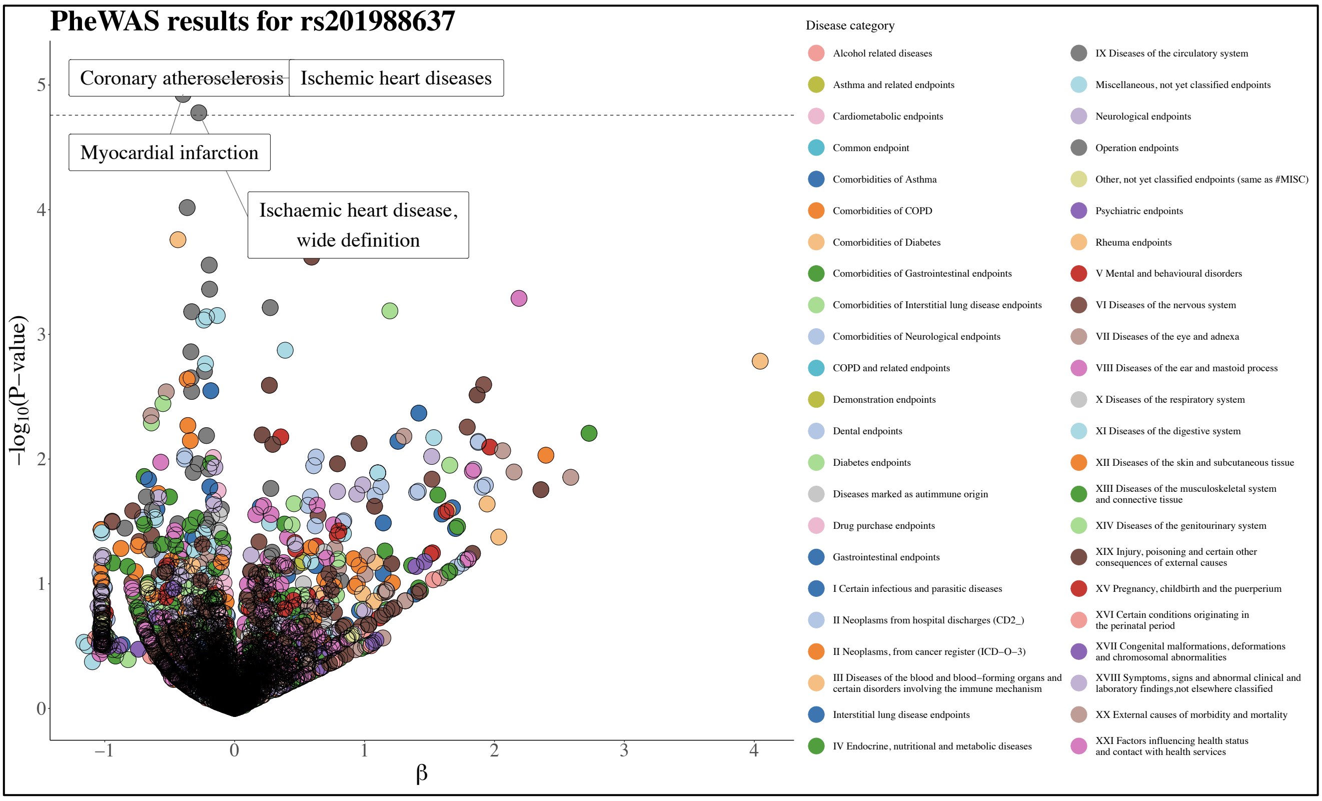


***Supplementary Figure 5:*** *Phenome-wide association study (PheWAS) results for rs201988637. Total number of endpoints is 2 861. Dashed line represents the phenome-wide significance threshold, multiple testing corrected by the number of endpoints = 0.05/ 2 861 =* $1.75\times10^{-5}$*. All endpoints reaching that threshold are labelled.*

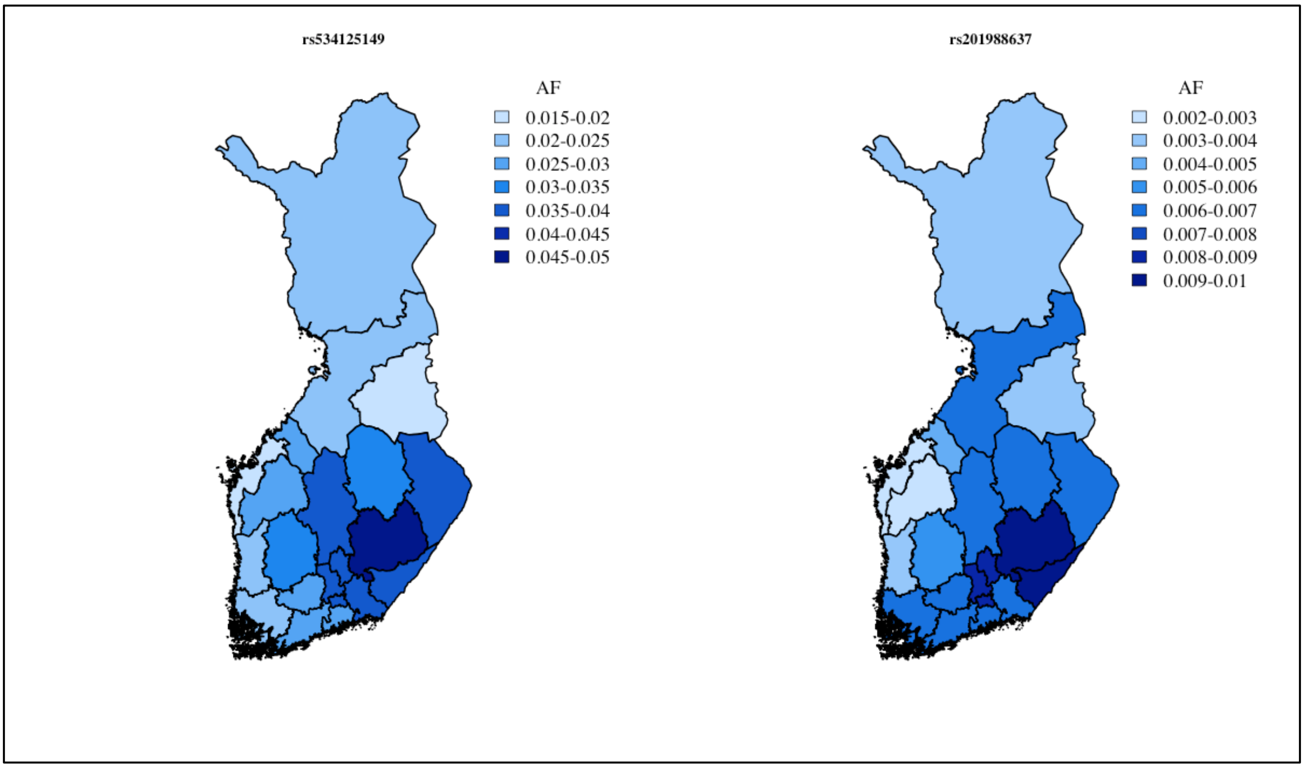


***Supplementary Figure 6:*** *Distribution of rs534125149 (inframe insertion) and rs201988637 (splice acceptor variant) across Finland.*

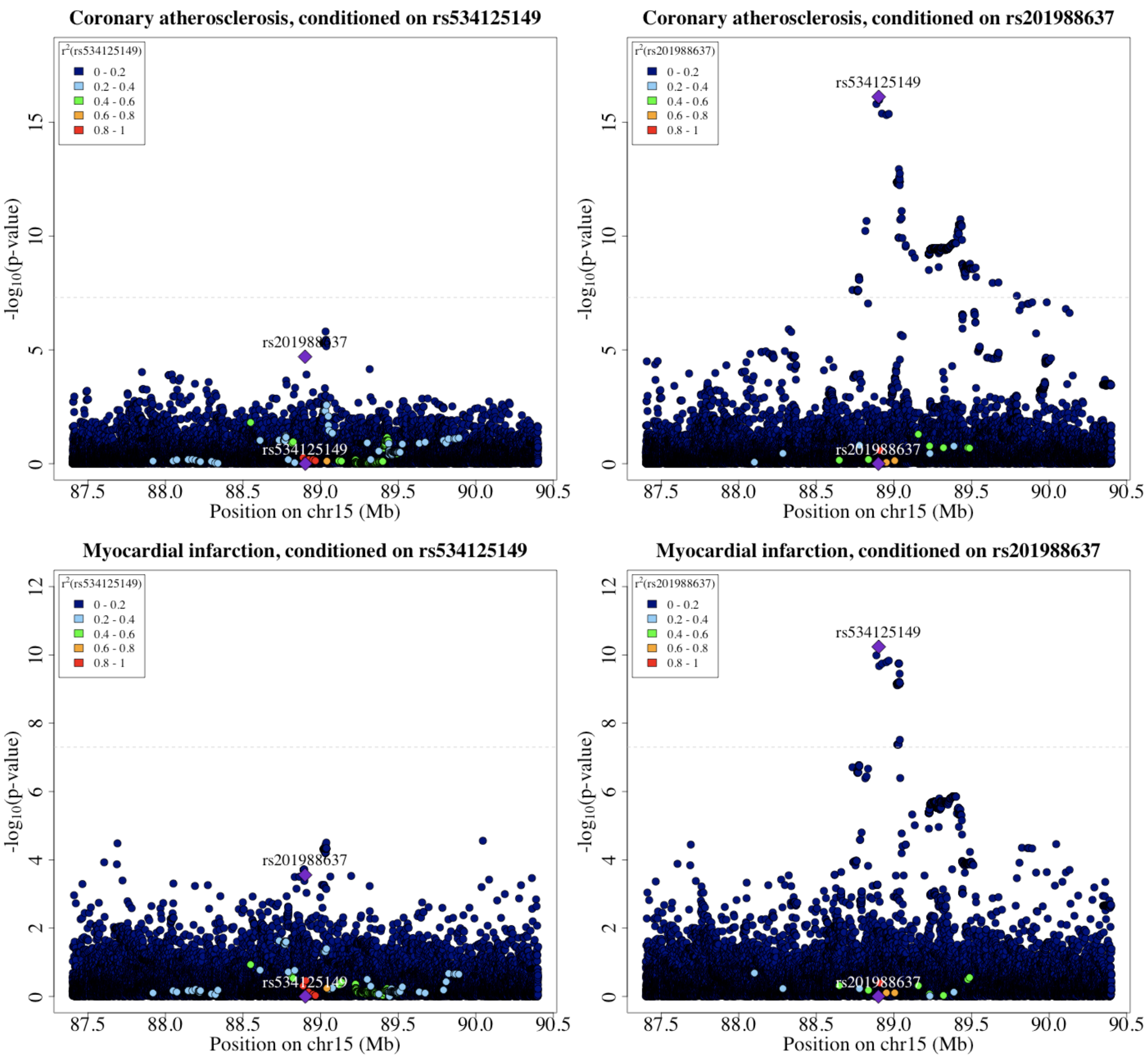


***Supplementary Figure 7:*** *Locuszoomplots from conditional analysis on rs534125149 and rs201988637 for coronary atherosclerosis and myocardial infarction (strict definition) on ±1.5 Mb region around rs534125149.*

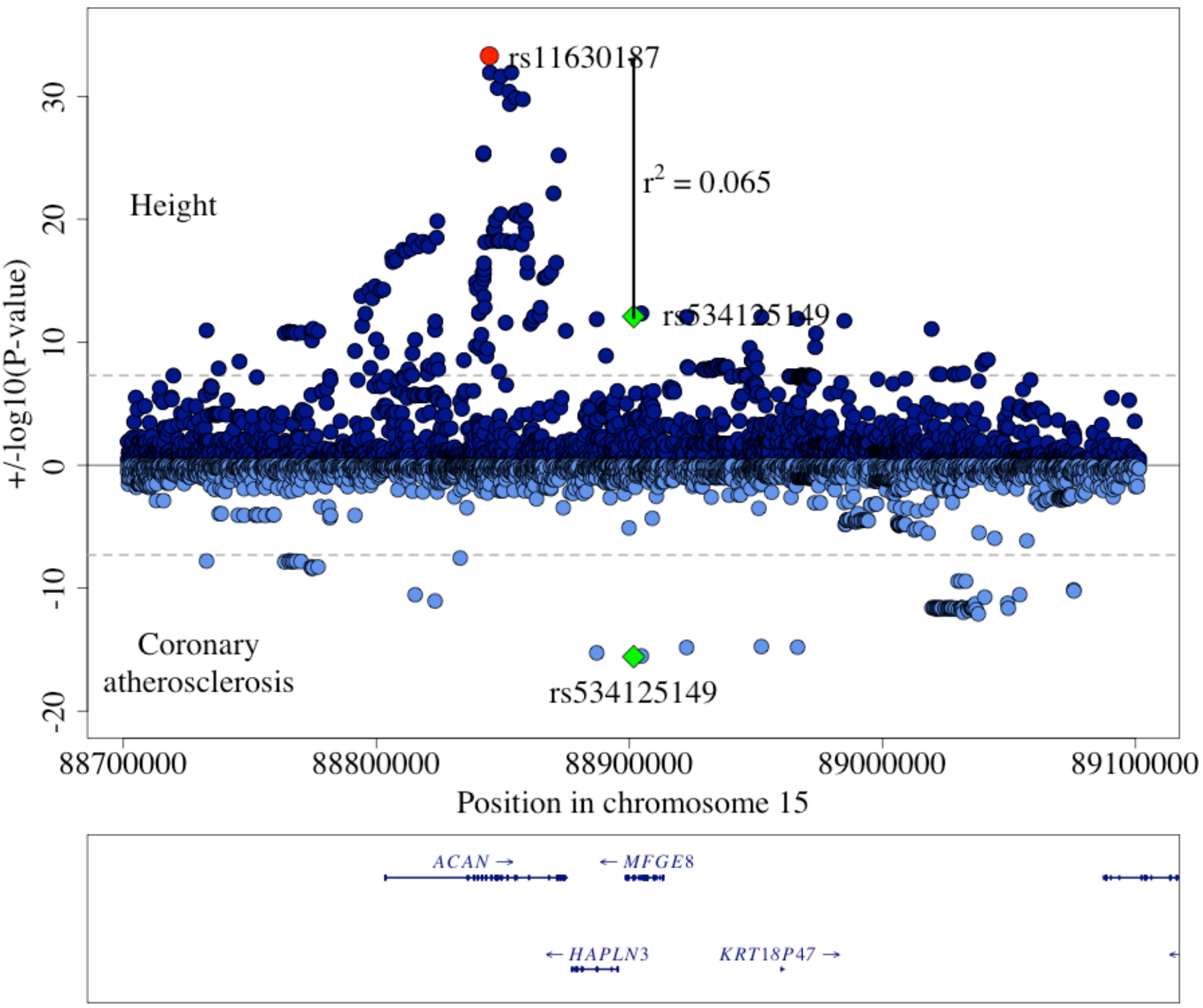


***Supplementary Figure 8:*** *Locuszoomplot for height and coronary atherosclerosis in ±200 kb region around rs534125149.*

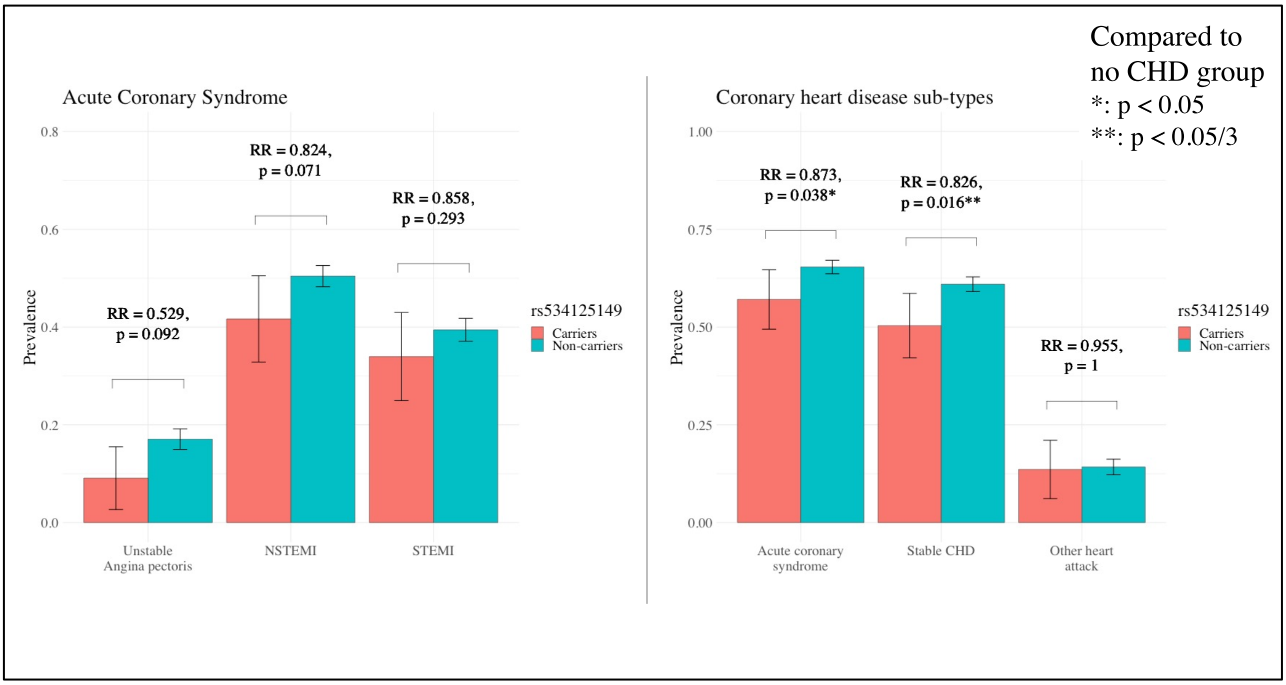


***Supplementary Figure 9:*** *Association of rs534125149 with sub-types of coronary heart disease and acute coronary syndrome. Comparison group is always no-CHD group.*

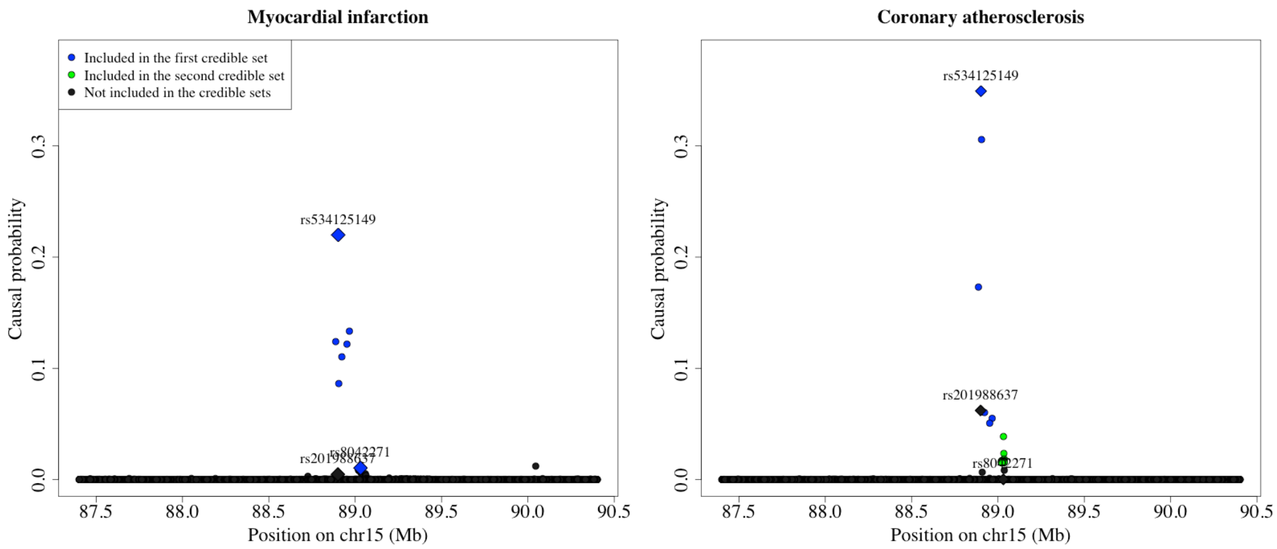


***Supplementary Figure 10:*** *Locuszoomplots from FINEMAP analyses for myocardial infarction and coronary atherosclerosis*.

### Supplementary tables

***Supplementary Table 1:*** *Significant association results from the phenome-wide scan.*

| Disease endpoint | Cases / Controls | rs534125149 (inframe insertion) | | rs201988637 (splice acceptor) | |
|---|---|---|---|---|---|
| | | OR [95% CI] | P-value | OR [95% CI] | P-value |
| Coronary atherosclerosis | 28 598/ 222 551 | 0.75 [0.71-0.81] | $2.63\times10^{-16}$ | 0.72 [0.63-0.83] | $7.94\times10^{-06}$ |
| Coronary revascularization (ANGIO or CABG) | 14 741/ 222 551 | 0.69 [0.63-0.76] | $1.86\times10^{-15}$ | 0.72 [0.59-0.87] | $6.58\times10^{-04}$ |
| Ischemic heart disease, wide definition | 39 358/ 221 047 | 0.80 [0.75-0.84] | $3.32\times10^{-14}$ | 0.76 [0.67-0.86] | $1.67\times10^{-05}$ |
| Ischemic heart diseases | 37 854/ 222 551 | 0.80 [0.75-0.84] | $4.26\times10^{-14}$ | 0.75 [0.66-0.85] | $9.10\times10^{-06}$ |
| Angina pectoris | 21 944/ 222 551 | 0.77 [0.72-0.83] | $4.38\times10^{-12}$ | 0.81 [0.69-0.94] | 0.01 |
| Major coronary heart disease event | 25 707/ 234 698 | 0.79 [0.73-0.84] | $1.12\times10^{-11}$ | 0.79 [0.68-0.92] | $1.98\times10^{-03}$ |
| Myocardial infarction, strict (only main diagnoses accepted) | 14 305/ 222 551 | 0.74 [0.68-0.81] | $1.95\times10^{-11}$ | 0.69 [0.58-0.83] | $9.62\times10^{-05}$ |
| Coronary artery bypass grafting | 6 904/ 222 551 | 0.66 [0.59-0.75] | $4.27\times10^{-11}$ | 0.73 [0.57-0.93] | 0.01 |
| Coronary angiopasty | 9 519/ 222 551 | 0.73 [0.65-0.80] | $6.51\times10^{-10}$ | 0.72 [0.58-0.89] | $2.87\times10^{-03}$ |
| Myocardial infarction | 15 787/ 222 551 | 0.78 [0.72-0.85] | $4.39\times10^{-09}$ | 0.67 [0.56-0.80] | $1.19\times10^{-05}$ |
| Major coronary heart disease event excluding revascularizations | 21 441/ 234 495 | 0.80 [0.75-0.87] | $5.43\times10^{-09}$ | 0.82 [0.70-0.96] | 0.01 |
| Hard cardiovascular diseases | 35 894/ 224 511 | 0.87 [0.82-0.92] | $1.06\times10^{-06}$ | 0.85 [0.75-0.96] | 0.01 |
| Status post acute MI | 1 881/ 222 551 | 0.61 [0.49-0.75] | $2.15\times10^{-06}$ | 0.65 [0.43-0.97] | 0.03 |
| Statin medication | 86 466/ 173 939 | 0.90 [0.86-0.94] | $1.54\times10^{-05}$ | 1.01 [0.91-1.12] | 0.82 |

*Association results for rs534125149 and rs201988637 for 14 endpoints with p-value < $1.75\times10^{-05}$ (PWS) for rs534125149.*

***Supplementary Table 2:*** *44 independent GWS associations for disease endpoint coronary atherosclerosis. Top variant of each independent loci represents one row.*

| SNPID (chr:pos:ref:alt) | rsid | Most severe consequence | Gene | CAD locus | Associated CAD-related trait(s) | FIN enrichment | AF | OR (P-value) |
|---|---|---|---|---|---|---|---|---|
| chr9:22099569:C:A | rs1537371 | Intron variant | *CDKN2B-AS1* | Yes | CAD, CHD | 0.8 | 0.42 | 1.25 ($2.30\times10^{-87}$) |
| chr6:12903725:A:G | rs9349379 | Intron variant | *PHACTR1* | Yes | CAD, CHD | 1.11 | 0.45 | 1.15 ($9.00\times10^{-33}$) |
| chr6:160589086:A:G | rs10455872 | Intron variant | *LPA* | Yes | Lp(a), CAD, LDL-C | 0.41 | 0.05 | 1.36 ($6.70\times10^{-30}$) |
| chr15:78750846:T:TAG | rs10625725 | Upstream gene variant | *ADAMTS7* | Yes | CHD, DBP, CAC | 0.842 | 0.64 | 0.9 ($5.10\times10^{-19}$) |
| chr1:55039974:G:T | rs11591147 | Missense variant | *PCSK9* | Yes | LDL-C, TC, CAD, MI | 3.1 | 0.04 | 0.77 ($4.10\times10^{-17}$) |
| chr19:11087511:C:T | rs17248720 | Upstream gene variant | *LDLR* | Yes | LDL-C, TC, CAD | 0.875 | 0.10 | 0.86 ($2.00\times10^{-16}$) |
| **chr15:88901702:C:CTGT** | **rs534125149** | **Inframe insertion** | ***MFGE8*** | **NO** | **-** | **inf** | **0.03** | **0.75 ($2.60\times10^{-16}$)** |
| chr4:147472512:C:T | rs1878406 | Intergenic variant | *EDNRA* | Yes | PP, CAD | 0.963 | 0.13 | 1.15 ($1.00\times10^{-15}$) |
| chr1:109275684:G:T | rs629301 | 3 prime UTR variant | *CELSR2* | Yes | LDL-C, TC, CAD | 0.989 | 0.79 | 1.12 ($1.60\times10^{-15}$) |
| chr19:44908822:C:T | rs7412 | Missense variant | *APOE* | Yes | CAD, LDL-C, TC | 0.561 | 0.05 | 0.83 ($4.20\times10^{-14}$) |
| chr14:99649135:C:T | rs541148696 | Intron variant | *HHIPL1* | Yes | CAD, SBP, MI | 175 | 0.01 | 1.51 ($9.80\times10^{-14}$) |
| chr21:34221526:G:A | rs28451064 | Intron variant | *AP000317.1/2* | Yes | CAD | 1.14 | 0.15 | 1.12 (1.20x10-13) |
| chr11:116752497:TA:T | rs66505542, rs398017699 | Intron variant | *BUD13* | Yes | TG, HDL-C, TC | 0.97 | 0.82 | 0.9 ($4.80\times10^{-13}$) |
| chr13:110397276:T:C | rs9515203 | Intron variant | *COL4A2* | Yes | CAD, LDL-C, | 1.01 | 0.26 | 0.91 ($2.50\times10^{-12}$) |
| chr12:111569952:C:T | rs653178 | Intron variant | *ATXN2* | Yes | BP, LDL-C, CAD | 1.16 | 0.58 | 0.92 ($3.40\times10^{-12}$) |
| chr1:56506681:G:A | rs72664324 | Intron variant | *PLPP3* | Yes | CAD, CHD, PP | 1.18 | 0.11 | 0.88 ($7.30\times10^{-12}$) |
| chr2:203206233:A:G | rs72936353 | Intron variant | *NBEAL1* | Yes | CAD | 0.803 | 0.11 | 1.13 ($3.40\times10^{-11}$) |
| chr2:85532371:C:T | rs59877521 | Intergenic variant | *MAT2A* | Yes | CAD | 1.03 | 0.43 | 1.08 ($1.30\times10^{-10}$) |
| chr7:150993088:C:T | rs3918226 | Intron variant | *NOS3* | Yes | CAD | 0.916 | 0.07 | 1.15 ($1.70\times10^{-10}$) |
| chr7:106770331:G:A | rs12705390 | Upstream gene variant | *PIK3CG* | Yes | TG | 1.54 | 0.30 | 1.08 ($3.00\times10^{-10}$) |
| chr17:2264939:C:T | rs7209564 | Intron variant | *SMG6* | Yes | CHD, SBP, BMI | 0.973 | 0.72 | 1.08 ($4.00\times10^{-10}$) |
| chr1:37950638:C:T | rs72661887 | Upstream gene variant | *INPP5B* | Yes | SBP, PP | 1.24 | 0.54 | 1.07 ($6.00\times10^{-10}$) |
| chr3:138369222:G:C | rs1199337 | Intron variant | *MRAS* | Yes | CAFD CHD, BMI | 1 | 0.14 | 1.11 ($6.80\times10^{-10}$) |
| chr20:45910063:C:A | rs201557719 | Missense variant | *PLTP* | Yes | HDL-C, TG | inf | 0.00 | 2.01 ($1.40\times10^{-09}$) |
| **chr6:130483492:A:G** | **rs118042209** | **Intergenic variant** | ***TMEM200A*** | **NO** | **-** | **0.79** | **0.01** | **0.7 ($1.90\times10^{-09}$)** |
| chr7:19013110:G:A | rs57301765 | Intergenic variant | *TWIST1* | Yes | PP, CAD, SBP | 1.3 | 0.19 | 1.09 ($2.10\times10^{-09}$) |
| chr12:54120216:A:T | rs11170821 | Non coding transcript exon variant | *SMUG1* | Yes | CAD, BMI | 2.21 | 0.06 | 1.15 ($2.40\times10^{-09}$) |
| chr5:108716278:T:C | rs6867978 | Intergenic variant | *FER* | Yes | SBP, PP | 0.854 | 0.29 | 0.93 ($2.80\times10^{-09}$) |
| chr12:57146968:C:G | rs7968719 | Intron variant | *LRP1* | Yes | CAD, BMI | 0.991 | 0.52 | 1.07 ($3.50\times10^{-09}$) |
| chr6:32204288:T:G | rs1044506 | Synonymous variant | *NOTCH4* | Yes | BMI | 1.01 | 0.87 | 1.1 ($6.80\times10^{-09}$) |
| chr6:133696717:G:A | rs34537042 | Intron variant | *EYA4* | Yes | TG, AF, WHR | 0.938 | 0.18 | 1.09 ($7.50\times10^{-09}$) |
| chr15:85610093:A:G | rs16942194 | Intron variant | *AKAP13* | Yes | SBP, PP, CAD | 2.09 | 0.10 | 0.9 ($8.10\times10^{-09}$) |
| chr4:41669720:A:T | rs141987800 | Intron variant | *LIMCH1* | Yes | LDL-C, Statin response | 0.52 | 0.00 | 1.75 ($9.40\times10^{-09}$) |
| chr8:22172265:C:A | rs73225841 | Intron variant | *BMP1* | Yes | CAD | 2.65 | 0.15 | 1.1 ($1.20\times10^{-08}$) |
| chr2:43845437:G:T | rs4299376 | Intron variant | *ABCG8* | Yes | LDL-C, TC, CAD | 1.16 | 0.78 | 0.93 ($1.70\times10^{-08}$) |
| **chrX:136194941:C:G** | **rs5974585** | **Intron variant** | ***FHL1*** | **NO** | **-** | **1.25** | **099** | **0.95 ($2.55\times10^{-08}$)** |
| chr8:125487789:C:G | rs28601761 | Intron variant | *TRIB1* | Yes | TG, HDL-C, MI | 1 | 0.41 | 0.94 ($2.60\times10^{-08}$) |
| **chr11:57380633:A:G** | **rs764568652** | **Intron variant** | ***PRG3*** | **NO** | **-** | **inf** | 0.00 | 7.72 ($4.10\times10^{-08}$) |

**Supplementary Table 3:** Characteristics of variants in MFGE8.

| Chr | Position[a] | Alleles (Ref/Alt) | rsid | Af (alt) | Info | Most severe consequence | ld with rs534125149 | ld with rs201988637 | ld with rs8042271 |
|---|---|---|---|---|---|---|---|---|---|
| 15 | 88901702 | C/CTGT | rs534125149 | 0.029 | 0.990 | Inframe insertion | 1 | 0.0001 | 0.1536 |
| 15 | 88899813 | T/G | rs201988637 | 0.006 | 0.986 | Splice acceptor variant | 0.0001 | 1 | 0.0010 |
| 15 | 89030987 | G/A | rs8042271 | 0.148 | 0.988 | Intergenic variant | 0.1536 | 0.0010 | 1 |

[a] *Positions are in build 38*

***Supplementary Table 4:*** *95% credible set for myocardial infarction (strict definition) from FINEMAP. Inframe insertion (rs534125149) and previously known common variant (rs8042271) are bolded.*

| SNPID (chr:pos:ref:alt) | rsid | Causal probability | ld with rs534125149 |
|---|---|---|---|
| **chr15:88901702:C:CTGT** | **rs534125149** | **0.250** | **1.000** |
| chr15:88966448:A:C | rs12594129 | 0.156 | 0.962 |
| chr15:88952146:C:T | rs150599388 | 0.141 | 0.963 |
| chr15:88887075:G:T | rs565898017 | 0.136 | 0.991 |
| chr15:88922596:C:T | rs117951873 | 0.125 | 0.967 |
| chr15:88904754:C:T | rs191156695 | 0.089 | 0.981 |
| **chr15:89030987:G:A** | **rs8042271** | **0.003** | **0.154** |
| chr15:89032865:T:A | rs56015348 | 0.003 | 0.154 |
| chr15:89029825:A:C | rs2003967 | 0.003 | 0.154 |
| chr15:89037068:C:G | rs2351254 | 0.002 | 0.112 |
| chr15:89034032:T:C | rs8035408 | 0.002 | 0.112 |
| chr15:89036690:G:C | rs7164299 | 0.002 | 0.114 |
| chr15:89026237:T:G | rs4932463 | 0.002 | 0.112 |
| chr15:89026272:G:C | rs4932208 | 0.002 | 0.112 |
| chr15:89028938:C:T | rs1550476 | 0.002 | 0.112 |
| chr15:89022026:A:C | rs1807214 | 0.002 | 0.112 |
| chr15:89023347:T:C | rs730657 | 0.002 | 0.112 |
| chr15:89024863:C:A | rs34166180 | 0.002 | 0.112 |
| chr15:89030984:A:G | rs8023801 | 0.002 | 0.112 |
| chr15:89031951:T:C | rs7162318 | 0.002 | 0.112 |
| chr15:89033730:T:C | rs28547445 | 0.002 | 0.112 |
| chr15:89021271:A:T | rs12592098 | 0.002 | 0.112 |
| chr15:89022834:C:T | rs735836 | 0.002 | 0.112 |
| chr15:89022640:T:A | rs735837 | 0.002 | 0.112 |
| chr15:89024874:A:AAAG | rs10683082 | 0.002 | 0.112 |
| chr15:89024799:T:G | rs9672658 | 0.002 | 0.112 |
| chr15:89026138:A:C | rs4932207 | 0.002 | 0.112 |
| chr15:89027167:G:C | rs28737395 | 0.002 | 0.112 |
| chr15:89026859:A:C | rs8041298 | 0.002 | 0.112 |
| chr15:89027456:A:G | rs12437794 | 0.002 | 0.112 |
| chr15:89027517:T:C | rs7169670 | 0.002 | 0.112 |
| chr15:89027435:G:C | rs7162610 | 0.002 | 0.112 |

***Supplementary Table 5****: 95% credible sets for coronary atherosclerosis. Inframe insertion variant (rs534125149) and bolded.*

| Credible set 1 | | | | Credible set 2 | | | |
|---|---|---|---|---|---|---|---|
| **SNPID chr:pos:ref:alt** | **rsid** | **Causal prob** | **ld with rs534125149** | **SNPID chr:pos:ref:alt** | **rsid** | **Causal prob** | **ld with rs7162326** |
| **chr15:88901702:C:CTGT** | **rs534125149** | **0.318** | **1.000** | chr15:89031969:T:C | rs7162326 | 0.048 | 1.000 |
| chr15:88904754:C:T | rs191156695 | 0.309 | 0.981 | chr15:89034032:T:C | rs8035408 | 0.028 | 0.952 |
| chr15:88887075:G:T | rs565898017 | 0.184 | 0.991 | chr15:89037068:C:G | rs2351254 | 0.025 | 0.950 |
| chr15:88922596:C:T | rs117951873 | 0.064 | 0.967 | chr15:89033989:G:A | rs8029102 | 0.021 | 0.952 |
| chr15:88966448:A:C | rs12594129 | 0.059 | 0.962 | chr15:89030984:A:G | rs8023801 | 0.021 | 0.952 |
| chr15:88952146:C:T | rs150599388 | 0.055 | 0.963 | chr15:89031951:T:C | rs7162318 | 0.021 | 0.952 |
| | | | | chr15:89035227:A:T | rs8037109 | 0.021 | 0.952 |
| | | | | chr15:89033730:T:C | rs28547445 | 0.021 | 0.952 |
| | | | | chr15:89031195:C:G | rs2083459 | 0.021 | 0.952 |
| | | | | chr15:89031187:T:C | rs2083458 | 0.021 | 0.952 |
| | | | | chr15:89026237:T:G | rs4932463 | 0.021 | 0.952 |
| | | | | chr15:89026272:G:C | rs4932208 | 0.021 | 0.952 |
| | | | | chr15:89028938:C:T | rs1550476 | 0.021 | 0.952 |
| | | | | chr15:89025698:A:G | rs4932206 | 0.021 | 0.952 |
| | | | | chr15:89035651:A:G | rs7182265 | 0.021 | 0.952 |
| | | | | chr15:89035631:A:T | rs7182131 | 0.021 | 0.952 |
| | | | | chr15:89031158:A:G | rs2083457 | 0.021 | 0.952 |
| | | | | chr15:89031253:T:C | rs2083460 | 0.021 | 0.952 |
| | | | | chr15:89031209:GT:G | rs35590540 | 0.021 | 0.952 |
| | | | | chr15:89031714:T:TG | rs35681620/ rs397714366 | 0.021 | 0.952 |
| | | | | chr15:89026138:A:C | rs4932207 | 0.021 | 0.952 |
| | | | | chr15:89035106:T:C | rs8041247 | 0.021 | 0.952 |
| | | | | chr15:89035116:G:C | rs8035119 | 0.021 | 0.952 |
| | | | | chr15:89035492:T:C | rs6496549 | 0.021 | 0.952 |
| | | | | chr15:89024863:C:A | rs34166180 | 0.021 | 0.952 |
| | | | | chr15:89036232:A:G | rs12593383 | 0.021 | 0.952 |
| | | | | chr15:89035688:A:T | rs7182288 | 0.021 | 0.952 |
| | | | | chr15:89024874:A:AAAG | rs10683082 | 0.021 | 0.952 |
| | | | | chr15:89024799:T:G | rs9672658 | 0.021 | 0.952 |
| | | | | chr15:89027167:G:C | rs28737395 | 0.021 | 0.952 |
| | | | | chr15:89026859:A:C | rs8041298 | 0.021 | 0.952 |
| | | | | chr15:89027456:A:G | rs12437794 | 0.021 | 0.952 |
| | | | | chr15:89027435:G:C | rs7162610 | 0.021 | 0.952 |
| | | | | chr15:89027517:T:C | rs7169670 | 0.021 | 0.952 |
| | | | | chr15:89021727:T:C | rs734780 | 0.021 | 0.952 |
| | | | | chr15:89022026:A:C | rs1807214 | 0.020 | 0.952 |
| | | | | chr15:89021271:A:T | rs12592098 | 0.020 | 0.952 |
| | | | | chr15:89022834:C:T | rs735836 | 0.020 | 0.952 |
| | | | | chr15:89022640:T:A | rs735837 | 0.020 | 0.952 |
| | | | | chr15:89023347:T:C | rs730657 | 0.020 | 0.952 |
| | | | | chr15:89022255:C:T | rs8037001 | 0.020 | 0.952 |
| | | | | chr15:89020702:T:C | rs28731436 | 0.020 | 0.952 |
| | | | | chr15:89020713:A:G | rs28735097 | 0.020 | 0.952 |
| | | | | chr15:89020231:C:G | rs1985691 | 0.020 | 0.952 |
| | | | | chr15:89019583:C:G | rs28687065 | 0.020 | 0.951 |

***Supplementary Table 6:*** *Association results for all 3 MFGE8 variants discussed for coronary atherosclerosis and MI, both separately and in the same model. Logistic regression adjusted for age, sex, genotyping batch and PC1-10 applied*

| Endpoint | rsid | SNPID (chr:position:ref:alt) | Separately | | All in the same model | |
|---|---|---|---|---|---|---|
| | | | OR [CI] | P | OR [CI] | P |
| Coronary atherosclerosis | rs534125149 | **chr15:88901702:C:CTGT** | **0.76 [0.71-0.81]** | **$8.30\times10^{-17}$** | **0.79 [0.74-0.85]** | **$1.14\times10^{-10}$** |
| | rs201988637 | chr15:88899813:T:G | 0.76 [0.66-0.87] | $1.04\times10^{-4}$ | 0.75 [0.65-0.86] | $4.41\times10^{-5}$ |
| | rs8042271 | chr15:89030987:G:A | 0.91 [0.88-0.94] | $1.49\times10^{-10}$ | 0.94 [0.91-0.97] | $4.56\times10^{-4}$ |
| Myocardial Infarction, strict | rs534125149 | **chr15:88901702:C:CTGT** | **0.74 [0.68-0.81]** | **$6.17\times10^{-11}$** | **0.79 [0.72-0.88]** | **$5.42\times10^{-6}$** |
| | rs201988637 | chr15:88899813:T:G | 0.72 [0.59-0.87] | $6.66\times10^{-4}$ | 0.70 [0.57-0.85] | $2.87\times10^{-4}$ |
| | rs8042271 | chr15:89030987:G:A | 0.88 [0.84-0.91] | $1.82\times10^{-10}$ | 0.91 [0.87-0.95] | $2.30\times10^{-5}$ |
